# High-resolution multiplexed antibody-omics and interpretable machine learning unveil novel pathogenic mechanisms in kidney transplant rejection

**DOI:** 10.1101/2025.07.25.25332230

**Authors:** Trirupa Chakraborty, Divya Bhakta, Anushka Saha, Camila Macedo, Daqiang Zhao, Asma Hashim, Kieran Manion, Marisa Abundis, Suhana Nujum Giyaz, Pedro Marcal, Alex Boshart, Aravind Cherukuri, Adriana Zeevi, Jeremy Tilstra, Alok Joglekar, Fadi Lakkis, Diana Metes, Ana Konvalinka, Aniruddh Sarkar, Jishnu Das

**Author notes:** Corresponding authors - Jishnu Das, Aniruddh Sarkar and Ana Konvalinka. These authors contributed equally (Divya Bhakta, Anushka Saha).

## Abstract

Antibody-mediated rejection (AbMR), driven by donor-specific alloantibodies (DSAs), is a major cause of late-stage kidney allograft failure, leading to premature graft loss in over half of affected patients. Despite efforts to link DSA features (e.g., HLA-specific IgG titers) to rejection risk, the immune mechanisms distinguishing DSA+ patients who develop AbMR remain unclear. In this first-in-class study, we develop a sample-sparing and cost-effective technique to generate the most comprehensive biophysical profile of DSAs reported to date. Further, given the complex pathological context and heterogeneity of samples we use a novel interpretable machine learning algorithm to learn signatures reflecting putative causal mechanisms of transplant rejection. We identify distinct mechanistically informative signatures at early and late times post-transplant. These antibody signatures, reflecting both quality and quantity of the humoral response, successfully discriminate DSA+ patients with and without AbMR. In addition to recapitulating known features of AbMR, our analyses reveal a significant and previously underappreciated role for IgM responses and glycosylation patterns, including sialylation and galactosylation, in both early and late rejection. Our identified signatures hold across two independent and geographically distinct cohorts.

Leveraging biomedical and computational innovation, we resolve prior inconsistencies in the field by implementing an unbiased systems framework identifying biophysical trends. These trends include selective enrichment of class I HLA-specific IgM and class II HLA-specific IgG responses in late and early rejection, respectively, which were overlooked earlier due to assay and methodological limitations. Corresponding functional relevance of putative causal signatures is further supported by observations from a murine model of chronic rejection, where we observe a significant increase in serum IgM-DSA associated with high risk of rejection as compared to serum IgG-DSA, warranting further exploration into the role of IgM in AbMR. Finally, addressing the lack of a comprehensive approach for pre-diagnosis of late AbMR patients reflecting the complex pathology of late AbMR and heterogeneity of samples (with time post-transplant ranging from 1-10 years), we formulate a risk score from our signatures. This composite risk score, combining IgM and sialylation metrics robustly predicts late AbMR with high sensitivity and specificity, offering a clinically actionable tool for early risk stratification. Together, leveraging our innovative pipeline we show the distinct roles of antibody isotypes/subclasses and glycosylation in disease progression, with IgM and glycosylation signatures showing strong diagnostic and prognostic value. Ultimately, the modularity of approach establishes a generalizable framework for understanding a plethora of complex immune-mediated tissue injury contexts beyond kidney transplantation.

## Introduction

Kidney transplantation is the best treatment for end-stage kidney disease. However, kidney transplants often fail prematurely. Antibody-mediated rejection (AbMR) accounts for more than 50% of late-stage allograft loss in kidney transplant patients, posing a significant challenge. Known to be primarily mediated by donor-specific alloantibodies (DSAs), AbMR involves activation of complement cascade and triggers inflammatory responses such as pattern-recognition receptor signaling, monocyte and macrophage responses, NK cell activity, and cytokine release(*1*, *2*) . AbMR is a heterogeneous disease, with distinct manifestations and clinical phenotypes. While the Banff criteria describe histopathological signatures used to identify and grade complex phenotypes of AbMR, understanding the immunological mechanisms underlying such phenotypes remains a key challenge in the field. Furthermore, numerous studies have repeatedly highlighted the need to integrate key clinical features such as the timing and extent of humoral responses, DSA profiles or molecular markers(*3*), necessitating deeper profiles of complex AbMR phenotypes that capture underlying mechanisms and provide a rigorous framework for diagnosis and treatment.

Despite several studies suggesting that functional properties of DSAs are associated with AbMR, why only about half of DSA positive (DSA+) patients go on to develop allograft rejection remains one of the longest-standing questions in the field. The clinical presentation of AbMR (in DSA+ patients) largely depends on the timing of the rejection post-transplantation, with different treatment responses. The first category of rejectors is those with pre-existing DSA who develop rejection within the first couple of months post-transplant (defined as early AbMR), whereas patients with *de novo* DSA tend to present with rejection later post-transplant (defined as late AbMR). It has been recognized in the literature that these two presentations are associated with very different outcomes. Discovering distinct mechanisms, clinical implications, treatment strategies for early versus late AbMR, and understanding the properties that distinguish DSA+ patients who will or will not develop rejection, is therefore crucial for improving graft survival and patient outcomes.

To date, much of the understanding of the association of DSAs with AbMR outcomes has relied on an approximate measure of magnitude of one dimension of the humoral response, specifically IgG antibodies, that is clinically commonly used. This has been made possible using single-antigen bead technology that involves measuring DSAs bound to specific antigens coated on the surface of barcoded microbeads, allowing detection of antigen-specific DSAs with high specificity. An approximate quantitative estimate of IgG DSAs is made using a single median fluorescence intensity (MFI) metric of binding measured in undiluted serum, which is then often binarized with a set cutoff for clinical decision making. While this method does come with caveats(*4*, *5*) related to its actual quantitative ability, overall, the focus has largely been on the “quantity” of response driven by IgG and some of its subclasses(*6*). Critically, this completely overlooks the qualitative functional heterogeneity of DSAs driven by modifications in their Fc domains. In the past decade, we and others have demonstrated the need to move beyond antibody quantity alone to the measurement of a more comprehensive set of biophysical features for these antibodies (the “antibody-ome”), in broader contexts of infectious and parasitic disease (*7–10*). Recent studies (*11–14*) have implicated Fc domain modifications of DSAs, such as specific glycoforms (e.g. galactosylation, afucosylation), and the role of Fc-receptor binding, in predicting AbMR status and graft survival. However, such glycosylation studies in the AbMR context are often hindered by methodological limitations including the relatively low abundance yet high diversity of DSAs and high sample requirement and cost of techniques involved in traditional glycomics methods. Additionally, there are unanswered questions, such as the role of IgM anti-HLA-DSAs in predicting rejection events (*15–17*), the importance of class I as opposed to class II anti-HLAs DSAs (*18*), and the role of different glycosylation signatures (*19*). Taken together, there is a pressing need for a broader approach to investigating the humoral mechanisms underlying AbMR.

In this first-in-class study, we investigate a cohort of DSA+ patients who either developed AbMR or had no rejection on a protocol or clinically indicated allograft biopsy. We characterized anti-HLA antibodies in the sera collected at the time of the biopsy, using a highly multiplexed, pan-antigen approach. We developed and used a high-throughput, sample-sparing method for bead-based profiling of DSAs to record a wide array of biophysical properties, such as isotype/subclass, FcR-binding, C1q binding, and glycosylation signatures. Notably, to create highly resolved humoral profiles, we employed a pan-antigen approach comprising 128 antigens (HLA and non-HLA) (fig. S1A), thus generating the most comprehensive coupled Fab + Fc profile of DSAs reported to date. We then coupled this to SLIDE, an interpretable machine learning algorithm recently developed by us, to discover humoral signatures associated with rejection in DSA+ patients. SLIDE moves beyond the discovery of biomarkers to the inference of actual biological mechanisms and has been successfully used in a range of biological contexts (*10*, *20–23*). Our approach helped systematically characterize humoral profiles of rejection in DSA+ patients (both early and late) and uncover mechanistic insights into AbMR pathogenesis. We then validated the observations in an external cohort, uncovering significant differences between early and late responses, with a novel association of IgM-DSA with AbMR. Our findings also highlight a key role for glycosylation in distinguishing early and late rejectors, unveiling powerful multivariate signatures with potential therapeutic implications.

## Results

### Antibody-profiling coupled with interpretable machine learning (ML) to study putative causal signatures that differentiate DSA+ AbMR+ from DSA+ AbMR-patients

In a clinical setting, DSA specificities and IgG MFIs are monitored to assess risk of AbMR. Given that the existing gold standard in the field relies on measuring IgG levels, we first interrogated if DSA+ AbMR+ patients could be identified from a cohort of DSA+ AbMR-patients based on their IgG titers in serum, either for DSAs or anti-HLA Abs. We thus obtained sera, collected at the time of the biopsy, from DSA+ AbMR- and DSA+AbMR+ patients from Starzl Transplantation Institute at the University of Pittsburgh (Pitt cohort, supplementary table 1 and 2). Total IgG titers (measured by MFI) bound to antigen pools showed no significant difference between DSA+AbMR-(non-rejectors) and DSA+AbMR+ (rejectors) patients (fig. S2A). Next, we developed and used a more comprehensive Fc profiling strategy to attempt to better distinguish between the two groups. To capture the full spectrum of antibody responses, including their Fc diversity, toward HLA and a subset of non-HLA antigens, we systematically profiled biophysical properties of antibodies against a wide range of antigen specificities. Using a high-throughput bead-based platform, we measured an array of antibody properties—such as isotype/subclasses, Fc-receptor binding, C1q binding and lectin interaction based glycoprofiles—against pools of 128 HLA/non-HLA antigens, gathering a comprehensive humoral profile of DSA+AbMR- and DSA+AbMR+ patients’ sera (**Fig. 1A-B**). This strategy allowed us to capture both Fab and Fc responses in a sample-sparing and cost-effective manner. The antigen pools included Class I-HLA (CI.1-CI.12), Class II-HLA (CII.1-CII.5), and non-HLA antigens MICA I (MICA.1) and MICA II (MICA.2) (fig.S1.A, 19 minipools corresponding to 128 unique antigens), while the probes targeted antibody isotypes/subclasses (total IgG, IgG1, IgG3, IgM), complement-binding C1q, relevant Fc-receptors (FcR2B, FcR3A), as well as glycosylation measured via lectin-based glycoprofiling—sialylation (SNA) and galactosylation (RCA). Notably, prior work, using traditional glycomics methods (e.g. Mass Spectrometry (MS)) has been limited to measuring Fc properties, especially glycosylation, of one or two selected antigen-specificities of DSAs due to the need to isolate and elute each antigen-specific antibody separately, resulting in high sample requirement and cost. Here we harness a lectin-binding based glycoprofiling assay (*10*, *24*) that recapitulates selected glycan profiles of a large set of antigen-specificities in a multiplexed manner from <10µL of sample in a cost-effective manner. This generates coupled Fab + Fc profiles of a wide set of antigen-specific antibodies consisting of 171 antibody features (19 minipools x 9 probes = 171 features, an “antibody-omic profile”) for each patient. This is, to the best of our knowledge, the most comprehensive biophysical profile of DSAs measured to date.

**Figure 1.**
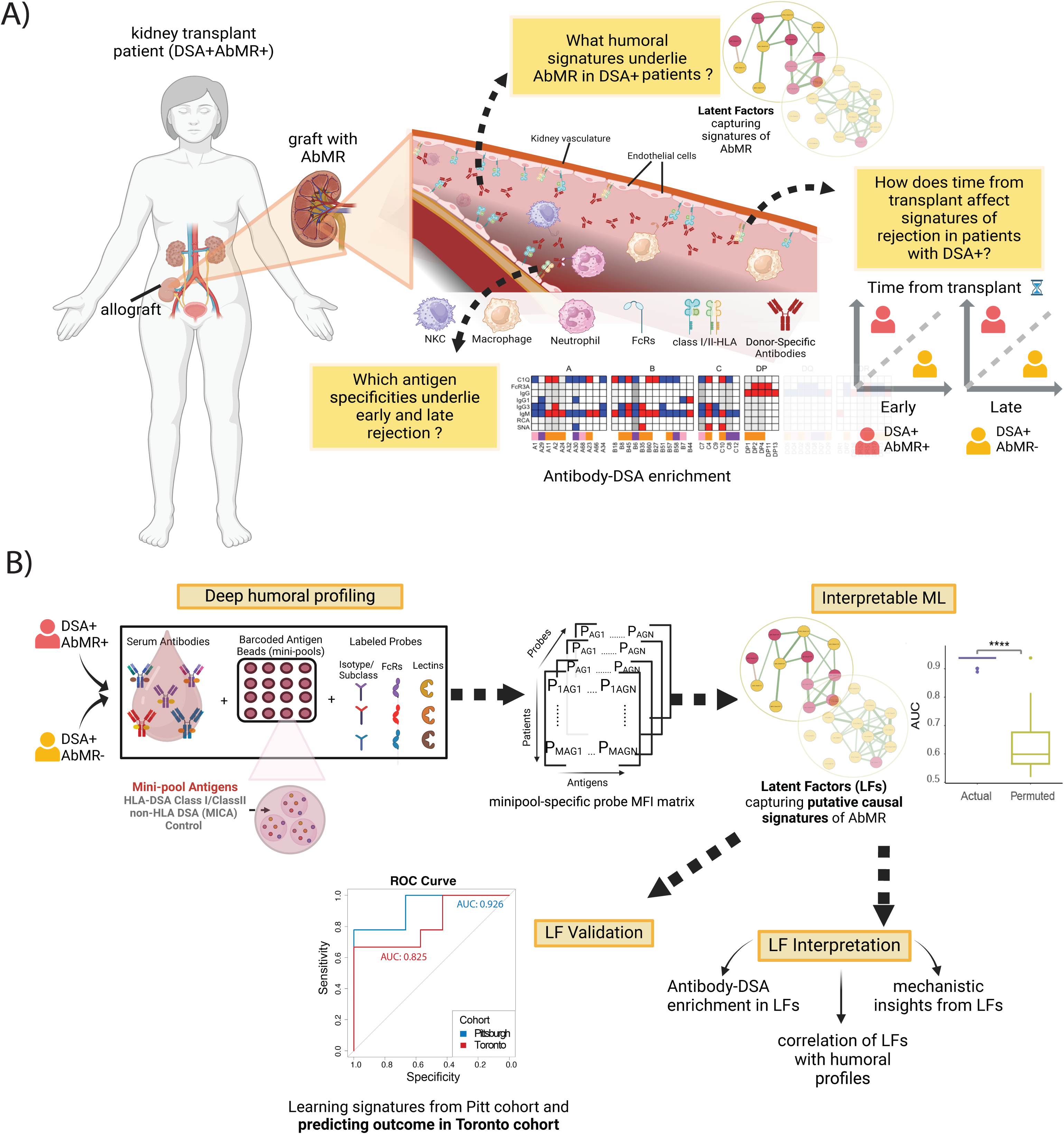
Approach and motivation of the study. A) Schematic of the broad questions addressed in the study. B) Schematic of the approach used in the study, coupling high-throughput humoral profiling with interpretable ML.

Previously, we and others have analyzed high-dimensional antibody-omic datasets using regularized regression (e.g. LASSO) and bootstrap aggregation in various disease contexts. However, these methods can only identify correlates of disease status and cannot infer molecular mechanisms of pathology. Here, we used SLIDE, an interpretable ML method recently developed by us (*20*) to move beyond predictive biomarkers to the actual inference of humoral signatures underlying pathogenesis. Built specifically for high dimensional data (low sample count with high features), SLIDE comes with rigorous statistical guarantees and identifies specific latent factors which are necessary and sufficient to infer differences between rejection outcomes (i.e., rejection or not) and are putatively causal. In contrast to clinical parameters which need a high sample size to show causation, by coupling SLIDE with humoral profiling, we can identify putative causal signatures giving deeper insights into AbMR pathogenesis. We have very recently shown that such an approach combining humoral profiling with interpretable ML can successfully identify humoral signatures underlying pathogenesis in the context of parasitic disease (*10*, *21–23*).Thus, here we implemented SLIDE to identify context-specific modules of humoral profiles that are necessary and sufficient to explain the differences between DSA+ patients who develop rejection (AbMR+) and those who do not (AbMR-).

### Elevated polyfunctional profiles characterize early time points of AbMR in DSA+ patients

We first looked at univariate measures of clinical outcomes (fig. S1B-F) to help describe significant differences in DSA+AbMR- and DSA+AbMR+ patients but being underpowered, they failed to provide deeper insights. Next, we used a more comprehensive approach using interpretable ML (SLIDE) to capture multivariate differences between the two groups that could have been missed by the univariate measures. However, the corresponding model still could not identify any significant differences in signatures between the DSA+AbMR+ and DSA+AbMR-groups (fig. S2B). This motivated us to ask if there were any clinical covariates that could better highlight possible differences between the two groups. Several studies in the field have demonstrated the importance of time from transplantation on graft survival, rejection outcomes and treatment response (*25*, *26*). To investigate if the time from kidney transplant could better inform humoral differences between the DSA+AbMR-group and the DSA+AbMR+ group, we stratified our cohort based on the median time from transplant (= 6 weeks for the Pittsburgh cohort), thus stratifying the patients into ‘early’ if they had a biopsy in the first 6 weeks post-transplant and ‘late’ if they had a biopsy after 6 weeks post-transplant. Unlike most clinical centers that only perform for-cause biopsies, more regular surveillance biopsies at Pittsburgh helped us converge on this unique truly early window and distinguish it from late rejection.

Humoral profiles of 18 patients who were categorized as ‘early’ (i.e. less than or equal to 6 weeks post-transplant, **Fig. 2A**) were then analyzed using SLIDE. The model identified 2 context-specific modules (the group-structure is inferred by SLIDE without prior knowledge) of humoral profiles (known as latent factors (LFs) - Z_E_1 and Z_E_2) that provided excellent discrimination between AbMR- and AbMR+ patients having DSAs in early stages post-transplant (model AUROC > 0.95 in a k-fold cross-validation framework, P < 0.05 with permutation testing) (**Fig. 2B**). The model utilized two latent factors; the first latent factor ( Z_E_1) (**Fig. 2C**, fig. S3A-I) comprises some known humoral profiles containing IgG1 and IgG3, which have previously been shown to be associated with AbMR (*6*, *12*, *13*), showing that the discriminatory signatures that our method identified, were able to recapitulate known correlates of rejection. The second latent factor (Z_E_2) contained predominantly sialylated antibodies against both class I and class II HLA antigens. Notably, both latent factors revealed signatures including elevated IgM and IgG antibodies against class I antigens in DSA+AbMR+ patients compared to DSA+AbMR-. While IgG subclasses like IgG1 and IgG3 have been associated with AbMR risk, the role of IgM in rejection has been unclear, at least in part because IgM antibodies are not routinely monitored. Interestingly, our unbiased machine learning approach identified modules indicating putative causal association of IgM against class I HLA with AbMR in DSA+ patients. The LFs also implicated an elevated sialylation (Sialic) profile associated with the DSA+AbMR+ group.

**Figure 2.**
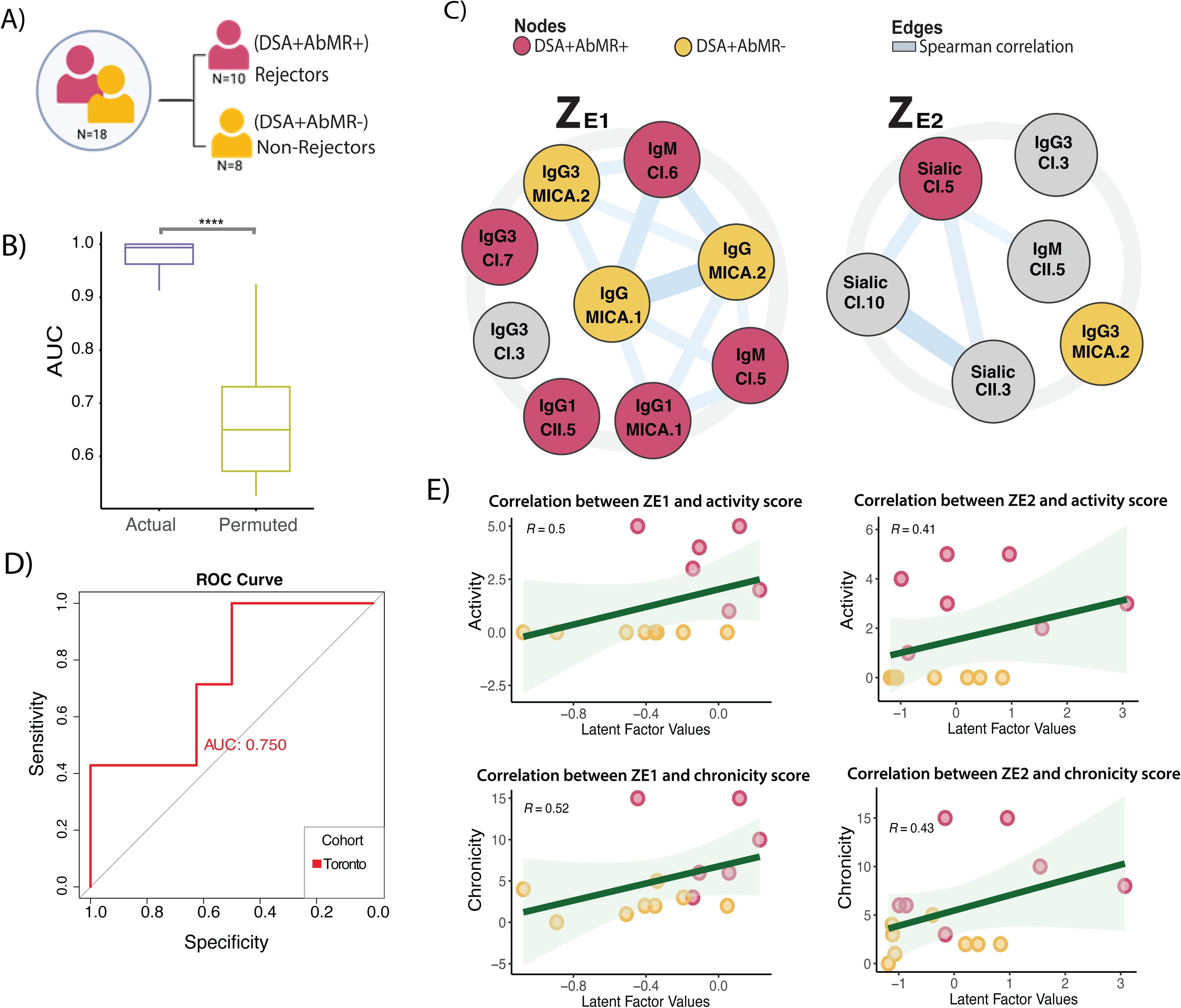
**Discovery and interpretation of humoral signatures that differentiate DSA+AbMR- from DSA+AbMR+ within the early group of patients.** A) Sample size of patients who were within 6 weeks from transplant, categorized as Early. The population of early patients (n=18) are further subdivided into DSA+AbMR+ (n=10) (Rejectors) and DSA+AbMR- (Non-rejector) patients (n=8). B) Performance of the real SLIDE model (in purple; AUROC ∼0.99) in discriminating between DSA+AbMR+ and DSA+AbMR-within the early group of samples using significant LFs, relative to the distribution of the performance of models built using permuted (shuffled) outcome labels (in green). Model performance was measured in a *k*-fold cross-validation framework with permutation testing, and the metric used is AUROC. **** indicates *P* < 0.0001 using the Mann-Whitney *U* test. C) Significant latent factors– Z_E_1 and Z_E_2, selected by SLIDE, discriminating between DSA+AbMR+ and DSA+AbMR-within the early group of samples. Larger circles in bold represent putative causal LFs. Each LF is visualized using a network representation. Edge thickness depicted with solid blue lines is based on absolute values of Spearman correlations; Nodes in red indicate humoral features higher in DSA+AbMR+ samples; Nodes in yellow indicate humoral features higher in DSA+AbMR-samples. Nodes in gray are unchanged between the two groups. Within each node, the corresponding feature includes the probe (in bold) with the corresponding bound minipool. CI = indicates a minipool with class I antigens, CII = indicates a minipool with class II antigens. D) ROCs showing SLIDE model performance using significant LFs in (C) to discriminate between DSA+AbMR+ and DSA+AbMR-patients within the early group of samples in the Toronto validation cohort (in red, AUROC= 0.75). The median time from transplant used to stratify early patients was cohort specific. Scatter plots showing correlation between latent factor values and (E) activity and (F) chronicity scores. Activity = g+ptc+v+c4d, chronicity= 2 x cg+ci+ct+cv (g= glomerulitis, ptc= peritubular capillaritis, v= vascular rejection, c4d= complement deposition marker, cg= chronic glomerulopathy, ci= interstitial fibrosis, ct= tubular atrophy, cv= chronic vascular changes, where each metric was graded between 0-3 by a clinician for each sample). Red dots correspond to DSA+AbMR+ patients and yellow dots correspond to DSA+AbMR-patients. R= spearman’s correlation coefficient

To rigorously test the generalizability of our observations from the Pittsburgh cohort, we sought to evaluate how predictive the identified signatures were in a geographically distinct cohort of patient samples obtained from the University Health Network, University of Toronto (Toronto cohort). Using only the humoral latent factors prioritized by the model after training on the Pittsburgh (discovery) cohort (i.e. using the humoral profiles picked up by Z_E_1 and Z_E_2), we assessed prediction performance on the validation cohort (cross-prediction). Since this involves learning both the structure and weights of the humoral signatures from solely the discovery cohort (we use the exact same 2 LFs identified from the Pittsburgh cohort with weights learnt using the Pittsburgh cohort), this constitutes a rigorous evaluation of generalizability of the identified signatures. The model remained significantly predictive for the Toronto cohort (AUROC on Toronto cohort= 0.75, P < 0.05, **Fig. 2D**, fig S4A-I), demonstrating that the prioritized LFs are not only robust, but also sufficient to explain differences between DSA+AbMR+ and DSA+AbMR-patients across orthogonal cohorts.

In clinical settings, the severity of rejection is estimated using metrics like activity and chronicity indices (*27*) based on Banff schema. While activity score describes the extent of glomerulitis, peritubular capillaritis, arteritis and complement deposition the chronicity score determines the extent of irreversible structural damage by incorporating measures of interstitial fibrosis, tubular atrophy, chronic vasculopathy and chronic glomerulopathy. Further, higher chronicity scores have been shown to be associated with graft loss. Therefore, we asked if the LFs correlated with histopathological manifestations of rejection. Both LFs showed a positive correlation with the activity and chronicity (**Fig. 2E**), thereby providing a plausible humoral signature for well-established histopathological metrics. However, as the correlation is only partial, the LFs seem to capture mechanisms beyond conventionally defined phenotypes.

### Late AbMR (DSA+AbMR+) is dominated by an IgM-specific signature that differentiates it from DSA+AbMR-patients

Late AbMR is clinically more challenging to treat and has worse prognosis compared to early AbMR. Owing to a chronic and often silent progression of rejection events, early detection of late AbMR poses a significant hurdle in the field (*28*). Given the poor response to standard anti-rejection therapies (*25*) and no approved treatment regimen for late AbMR (*26*), there is a pressing need for better understanding of mechanisms of late rejection for prediction and therapeutic insights. Thus, we sought to first identify distinct humoral signatures between DSA+AbMR+ patients developing AbMR beyond 6 weeks post-transplant (i.e. late AbMR) and DSA+AbMR-patients (**Fig. 3A**). Using SLIDE, we identified 5 significant LFs that provided significant discrimination between the DSA+AbMR- and DSA+AbMR+ patients (AUROC = 0.78, P < 0.05, **Figs. 3B-C**, fig. S5A-I). Of these, Z_L_1 captured an IgM-centric response primarily against class I HLAs elevated in rejector patients (DSA+AbMR+) compared to the non-rejector group (DSA+AbMR-). Strikingly, Z_L_2 and Z_L_3 were dominated by a low C1q-specific response in the rejectors compared to non-rejectors, indicating that rejectors had higher serum C1q binding (thus low free serum C1q captured in the assay) when their biopsies were obtained late post-transplant. This was further corroborated by a significantly higher C4d score in DSA+AbMR+ patients compared to DSA+AbMR-patients (fig. S7A) (patients belonging to Pitt cohort 2 were used for this due to the low sample size in Pitt cohort 1) The method also picked up modules (Z_L_4 and Z_L_5) with elevated IgG (particularly against class II HLA) and sialylation signatures. This observation contrasts with recent findings that suggested that DSA-IgG sialylation is not associated with pathogenicity and AbMR outcome (*19*).

**Figure 3.**
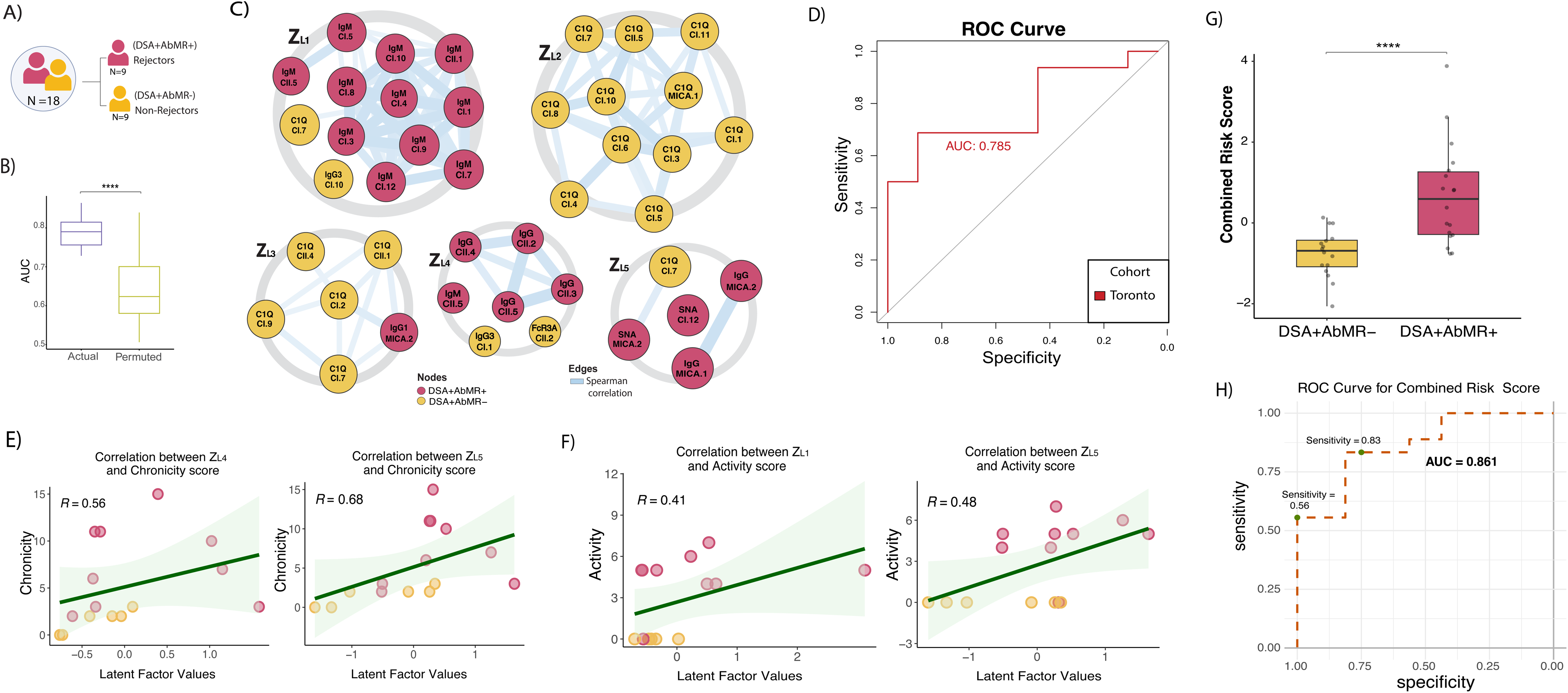
**Discovery and interpretation of humoral signatures that differentiate DSA+AbMR-from DSA+AbMR+ within the early group of patients.** A) Sample size of patients who were beyond 6 weeks from transplant, categorized as Late. The population of late patients (n=18) are further subdivided into DSA+AbMR+ (n=9) (Rejector) and DSA+AbMR-(Non-rejector) patients (n=9). B) Performance of the real SLIDE model (in purple; AUROC =0.93) in discriminating between DSA+AbMR+ and DSA+AbMR-within the late group of samples using significant LFs, relative to the distribution of the performance of models built using permuted (shuffled) outcome labels (in green). Model performance was measured in a *k*-fold cross-validation framework with permutation testing, and the metric used is AUROC. **** indicates *P* < 0.0001 using the Mann-Whitney *U* test. C) Significant latent factors– Z_L_1, Z_L_2, Z_L_3, Z_L_4 and Z_L_5, selected by SLIDE, discriminating between DSA+AbMR+ and DSA+AbMR-within the late group of samples. Larger circles in bold represent putative causal LFs. Each LF is visualized using a network representation. Edge thickness depicted with solid blue lines is based on absolute values of Spearman correlations; Nodes in red indicate humoral features higher in DSA+AbMR+ samples; Nodes in yellow indicate humoral features higher in DSA+AbMR-samples. Nodes in gray are unchanged between the two groups. Within each node, the corresponding feature includes the probe (in bold) with the corresponding bound minipool. CI = indicates a minipool with class I antigens, CII = indicates a minipool with class II antigens. D) ROCs showing SLIDE model performance using significant LFs in (C) to discriminate between DSA+AbMR+ and DSA+AbMR-patients within the late group of samples in the Toronto validation cohort (in red, AUROC= 0.78). The median time from transplant used to stratify late patients was based on the Pittsburgh cohort median. E) Scatter plots showing correlation between latent factor values and chronicity and scores and activity scores (F). chronicity= 2 x cg+ci+ct+cv (cg= chronic glomerulopathy, ci= interstitial fibrosis, ct= tubular atrophy, cv= chronic vascular changes, where each metric was graded between 0-3 by a clinician for each sample). Red dots correspond to DSA+AbMR+ patients and yellow dots correspond to DSA+AbMR-patients. R= spearman’s correlation coefficient. G) Boxplot showing Combined Risk Score values across pooled validation and discovery cohort samples, segregated into DSA+AbMR- and DSA+AbMR+ groups. **** indicates *P* < 0.0001 using the Mann-Whitney *U* test. H) ROC showing performance of the Combined Risk Score in predicting outcome (i.e. DSA+AbMR- or DSA+AbMR+) (AUROC= 0.86). Green dots correspond to sensitivity value at specificity of 1.00 (sensitivity= 0.56) and specificity of 0.75 (sensitivity = 0.83).

To evaluate the robustness of the multivariate signatures, we tested how well this late signature identified in the discovery (Pittsburgh) cohort was able to predict AbMR status in the validation (Toronto) cohort. Our analysis showed that the signatures captured by the 5 LFs were extremely predictive of rejection outcome in late time points of an orthogonal cohort (AUROC = 0.78 in the validation cohort, P < 0.05, **Fig. 3D**, fig. S6A-I), confirming the robustness of the identified signature. Of the 5 LFs identified by the model, we found that only Z_L_4 and Z_L_5 captured profiles strongly correlating with chronicity only (**Fig. 3E**), in contrast to the response in early time points, wherein both LFs showed a correlation with chronicity and activity. While Z_L_2, Z_L_3, Z_L_4 and Z_L_5 LFs seemed to explain/capture differences at the phenotypic level (C4d deposition (fig. S7A) and chronicity (**Fig. 3F**), we were intrigued that the IgM-dominant LF, Z_L_1 did not. Interestingly, elevated total IgG and sialylation signatures captured by Z_L_5 (sialylation LF) showed a strong correlation with chronicity. Overall, our results underscored a putative causal association of elevated IgM response with late AbMR within a group of DSA+ patients. Motivated by the strong dominance of the IgM and sialylation LFs, we looked at the distribution of samples (in both Pitt and Toronto cohorts) between the two LFs of interest and found that the DSA+AbMR-samples were largely negative for both LFs, compared to the DSA+AbMR+ (fig. S7B). Thus, we asked if a score combining the two signatures was sufficient to segregate between the rejector and non-rejector samples within the late cohort. This combined score, pooling multivariate signatures of IgM and sialylation LFs, showed a significant difference between rejector and non-rejector samples (**Fig. 3G**), across the discovery and validation cohorts. Further, the rejectors had an overall positive and higher score compared to the negative score in non-rejectors, thereby providing a clear humoral correlate of late AbMR risk. Further, we find that the combined risk score can predict late AbMR risk at performance of AUROC = 0.86 on samples across two geographically distinct cohorts (**Fig. 3H**), elucidating a novel humoral correlate of AbMR risk that can be clinically monitored. The inference of this intuitive novel metric was made possible because of the inherent interpretability of our machine learning approach (as opposed to black box techniques). Its performance in stratifying subjects with or without late AbMR across the Pitt and Toronto cohorts demonstrates its robustness.

### Early and late rejection profiles capture an enrichment in IgG1 against class II HLA and IgM against class I HLA

To further understand the humoral signatures capturing such previously under-appreciated antibody isotypes, we sought to inspect which antigenic specificities these biophysical features were enriched for. While the presence of preformed DSAs against class I and class II is a predictor of early AbMR (*29*), late AbMR has typically been associated with *de novo* DSAs against class II HLAs, particularly anti-DQ DSAs. These observations have been based on solid-phase assays, specifically, the single-antigen bead technology. Such an approach, focused only on IgG antibody measurements, neglects the specificities of other isotypes (and subclasses). It also fails to capture antibodies against other antigens such as non-HLA antigens, as well as antigens that are not donor-specific (non-DSA), that exhibit distinct functionality based on Fc receptor binding, glycosylation and ability to activate complement. In this study, we focused not just on DSAs against HLA molecules (including memory and *de novo* responses) but also examined antibodies against additional antigen specificities. To capture the breadth of humoral responses during early and late rejection in a high-throughput, multiplexed approach, we used HLA class-specific pools of antigen beads (referred to as mini pools, fig. S1A and **Fig. 4A**). Further, given the record of DSAs of patients in the Pittsburgh cohort, we were uniquely equipped to ask which DSAs / non-DSAs were more likely to be associated with early and late AbMR. Thus, we asked which anti-HLA DSA specificities were enriched in the humoral responses associated with early and late AbMR (**Fig. 4B**). We first stratified the tested antigens into three categories for early and late time subgroups respectively. The first were HLA antigens against which patients with AbMR developed DSAs (rejection-associated antigens), the second included HLA antigens against which patients with no rejection developed DSAs, and third were those HLA antigens against which there were no recorded DSAs at the time of or preceding the biopsy. Our analysis revealed that among DSA+ AbMR+ patients, IgG3 DSAs against class I HLAs (specifically the A68 serotype) were enriched. Similarly, among these DSA+ patients with AbMR, DSAs of the IgG1 subclass were predominantly directed against class II-HLA specificities including DR (DR52, DR53), DP (DP13) and DQ (DQ4 and DQ7) (**Fig. 4C**). Interestingly, in case of late AbMR we found a clear enrichment in IgM antibodies directed against donor class I HLA antigens in rejectors, as well as IgG DSAs against class II HLA antigens. In contrast, DSA against class I and class II HLA exhibited increased C1q binding in the non-rejectors (**Fig. 4D**). We found the same trend in enrichment in non-DSA anti-HLAs (fig. S8A-B). Such a serotype-specific association with AbMR on a global level was possible owing to the information on previous DSA serotypes that each patient developed across time and due to the power of our analytical pipeline.

**Figure 4.**
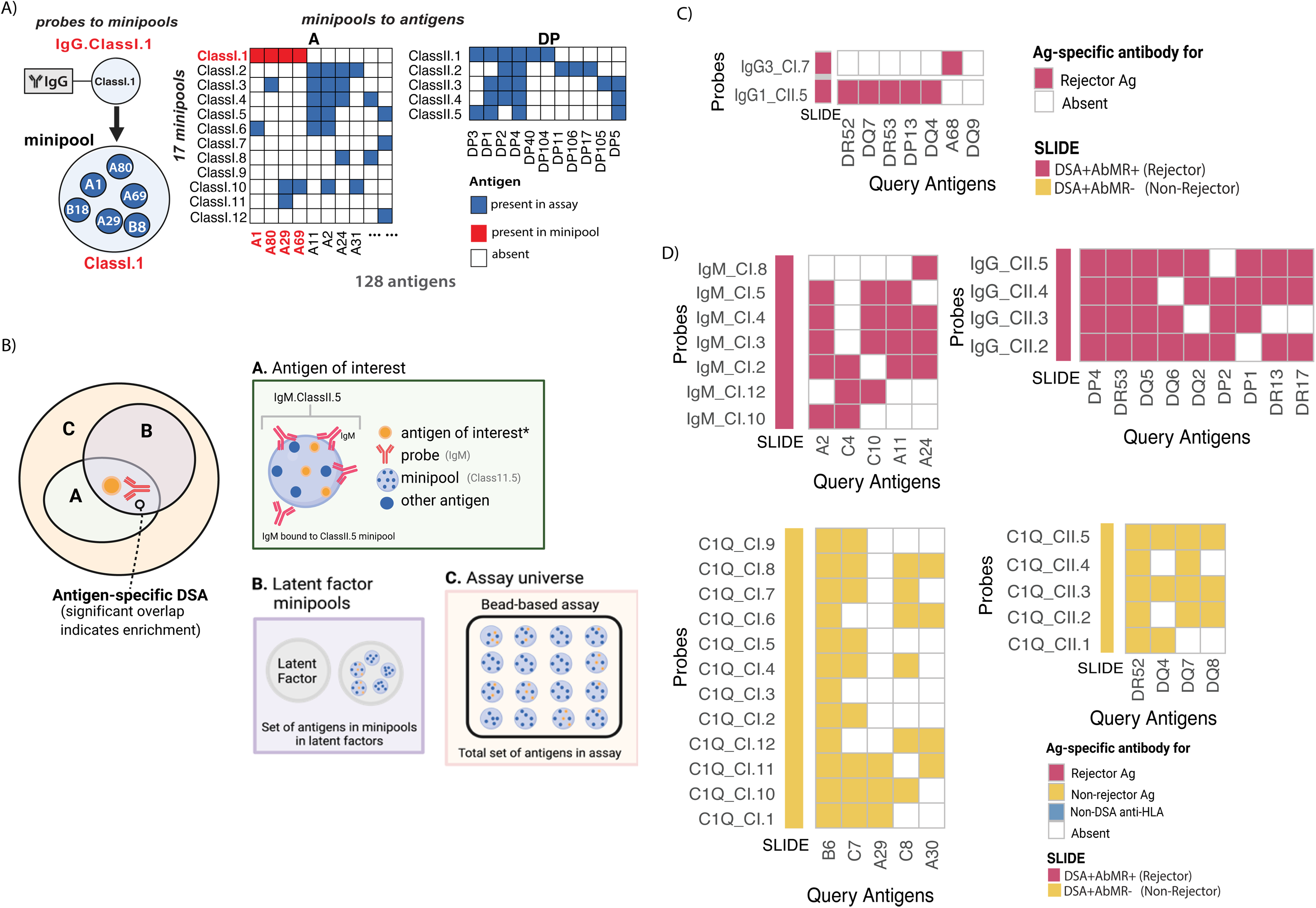
**SLIDE factors reveal an enrichment of specific DSA specificities in early and late rejection.** A) Schematic representation with an example showing mapping of minipool to set of pre-designed antigens, naming of minipools and the association with given probe. Antigens highlighted in blue are the set of antigens coated on minipool bead surface of the pool of antigens present in the experimental assay. (Only a truncated set of the total 128 antigens have been depicted here as an example). Serotypes of the corresponding antigens have been highlighted in red. B) Venn diagram depicting enrichment test for patient DSAs in latent factors where-A shows an example of a probe-minipool pair picked up by the latent factor with a bunch of antigens including the patient DSA of interest (query antigen) comprising a minipool; B shows example of minipools picked up by latent factors; C depicts the distribution of query antigen in the universe of antigens tested in the bead-based assay. Probe-minipool features selected by significant latent factors, enriched in patient DSAs (serotypes). Features that were elevated in DSA+AbMR+ (rejector) patients as compared to the DSA+AbMR-(non-rejector) patients in the latent factors from early (C) and late groups (D) respectively, are depicted in red in the SLIDE column, and those that were lower in early non-rejectors is depicted in yellow. Patient DSA serotypes that are in red are enriched in the rejectors for the corresponding probe (in the probe-minipool pair). Patient DSA serotypes that are in yellow are enriched in the non-rejectors for the corresponding probe (in the probe-minipool pair).

### Humoral signatures of early rejection can help predict late rejection

Given that distinct humoral signatures are associated with early and late rejection compared to the non-rejector group, we asked if early signatures could help predict outcomes in the late group. Our earlier analyses already identified a highly predictive composite risk score for late rejection using humoral profiles at the late timepoint. So, we turned to the next big challenge in the field - early prediction of late rejection. Using the significant LFs learned from the early model (**Fig. 2C**), we performed cross-predictions to predict outcomes at the late timepoint (**Fig. 5A(i)**) using humoral signatures learned at the early time point and vice-versa (**Fig. 5A(ii)**). Interestingly, we found that LFs learned from the early model (that discriminated between rejectors and non-rejectors) could successfully discriminate between rejectors and non-rejectors in the late group of patients (AUROC = 0.78) (**Fig. 5A(i)**). However, when we used significant LFs learned from the late model (**Fig. 3C**) to cross-predict outcomes in the early group of patients, the model prediction was not as good (AUROC = 0.66, **Fig. 5A(ii)**). These results indicate that early signatures can not only be used to predict which patients are likely to experience late rejection, implying shared humoral signatures between early and late rejection models. Juxtaposed with the correlations between LFs in early and late rejections with Banff activity and chronicity scores (in **Fig. 2E** and **Fig. 3E**), these demonstrate that early humoral responses include a chronicity signature which can be extrapolated to late events. However, the late signature is more nuanced and includes more than just a chronic signature, perhaps diverging further from early responses.

**Figure 5.**
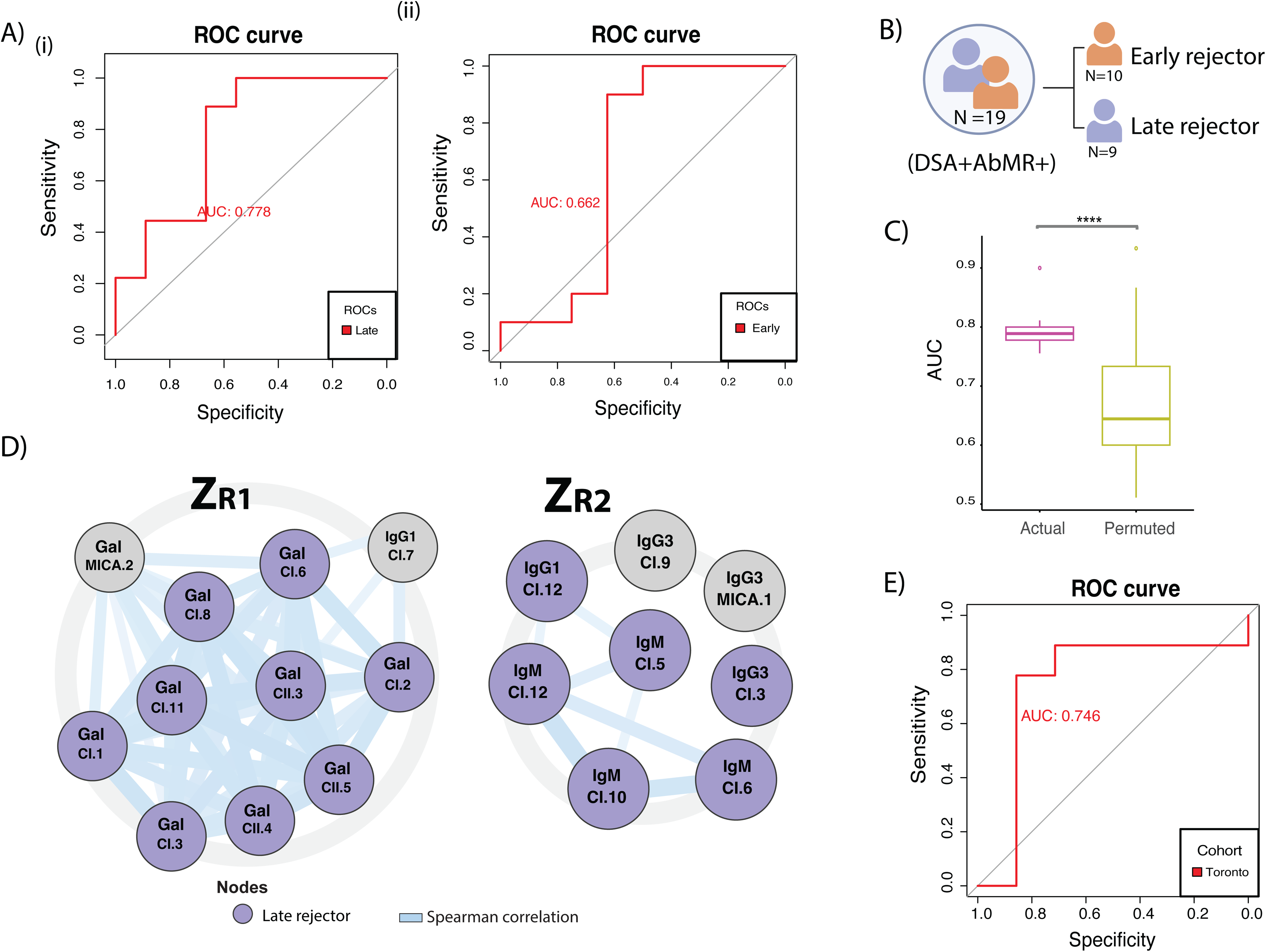
**Humoral signatures of early rejection can help predict late rejection outcomes.** A) (i) ROCs showing SLIDE performance using significant LFs obtained from Fig2.C (early model) and tested on the late group (late DSA+AbMR+ and late DSA+AbMR-) with AUROC = 0.78 (in red). (ii) ROCs showing SLIDE performance using significant LFs obtained from Fig3.C (late model) and tested on the early group (early DSA+AbMR+ and early DSA+AbMR-) with AUROC = 0.66 (in red). B) Sample size of patients who were categorized as early and late rejectors within the DSA+AbMR+ group in the full cohort. The population of rejector patients (n=19) are further subdivided into Early rejector (n=10) and Late rejector patients (n=9). C) Performance of the real SLIDE model (in purple; AUROC ∼0.79) in discriminating between early and late within the rejector group of samples using significant LFs, relative to the distribution of the performance of models built using permuted (shuffled) outcome labels (in green). Model performance was measured in a *k*-fold cross-validation framework with permutation testing, and the metric used is AUROC. **** indicates *P* < 0.0001 using the Mann-Whitney *U* test. D) Significant latent factors– Z_R_1, Z_R_2, selected by SLIDE, discriminating between early and late patients within the DSA+AbMR+ (rejector) group of samples. Larger circles in bold represent putative causal LFs. Each LF is visualized using a network representation. Edge thickness depicted with solid blue lines is based on absolute values of Spearman correlations; Nodes in purple indicate humoral features higher in late rejector samples; Nodes in orange indicate humoral features higher in early rejector samples. Nodes in gray are unchanged between the two groups. Within each node, the corresponding feature includes the probe (in bold) with the corresponding bound minipool. CI = indicates a minipool with class I antigens, CII = indicates a minipool with class II antigens. E) ROCs showing SLIDE model performance using significant LFs in (D) to discriminate between early and late rejector patients within the DSA+AbMR+ group of samples in the Toronto validation cohort (in red, AUROC= 0.75). The median time from transplant used to stratify early and late patients was cohort specific.

These findings motivated us to dissect humoral differences between early and late rejection. So, we used SLIDE to identify signatures that underlie differences between early and late rejector patients (**Fig. 5B)**. Our analysis revealed two LFs - Z_R_1 and Z_R_2 that captured significant differences between early and late rejection outcomes (**Fig. 5C**). Strikingly, the two LFs captured an overall elevated signature of galactosylation (Gal) and IgM in the late rejector group compared to the early rejectors (**Fig. 5D)**. While galactosylation of both antibodies directed against class I and class II HLA was increased in late rejectors, we found that the elevated IgM response was associated with antibodies against class I HLA antigen-pools in the late rejectors compared to the early. Further, using just the prioritized LFs learned from the Pittsburgh cohort the model was able to distinguish early and late rejection in the Toronto cohort (AUROC = 0.75, **Fig. 5E**), validating the robustness of the prioritized signatures. Taken together, our results highlight that while early responses typically comprise C1q-binding isotype/subclasses, the relative importance of IgM and glycosylation (galactosylation) are more pronounced at later time points, exposing novel pathways of understanding late rejection. Thus, our results strongly imply the need to clinically monitor these signatures for early detection of late AbMR.

### Serum DSA-IgM but not DSA-IgG is significantly elevated in murine models of high risk chronic rejection

Piqued by the core IgM signature that were the defining feature of the humoral signatures in late rejection, we asked if the same trends could be observed in a previously well-established murine model of rejection. Specifically, we sought to compare the levels of serum DSA-IgM in a murine model that best recapitulates late rejection - C57BL/6 mice receiving kidneys transplanted from NOD donors (high-risk) and C57BL/6 recipient mice with kidneys from NOD.B6 Sirpa donors (models mismatched MHC, but donor and recipient are matched for Sirpa) (**Fig. 6A**). Since Sirpa mismatch has been implicated in interstitial inflammation and peritubular capillaritis and graft failure(*27*), matched Sirpa in this model was associated with a lower risk of rejection. Interestingly, comparing serum MFIs of DSA-IgG across the two mice models at different time points - day 7, day 21 and day 360 of transplant (**Fig. 6B**), revealed no significant changes in serum DSA-IgG levels. However, strikingly, the high-risk model of chronic rejection (C57BL/6 recipient with kidney from NOD donor) had significantly higher serum DSA-IgM compared to the low-risk group, at days 7, 21 and 360 (**Fig. 6C**). Together, these results provide further support for our finding that serum DSA-IgM is potentially a strong indicator of AbMR risk. In current clinical settings, only serum DSA-IgG is measured. However, our findings from both the antibody-omic analyses coupled to machine learning and the murine model, suggest that serum DSA-IgM should be measured as it is informative of AbMR risk.

**Figure 6.**
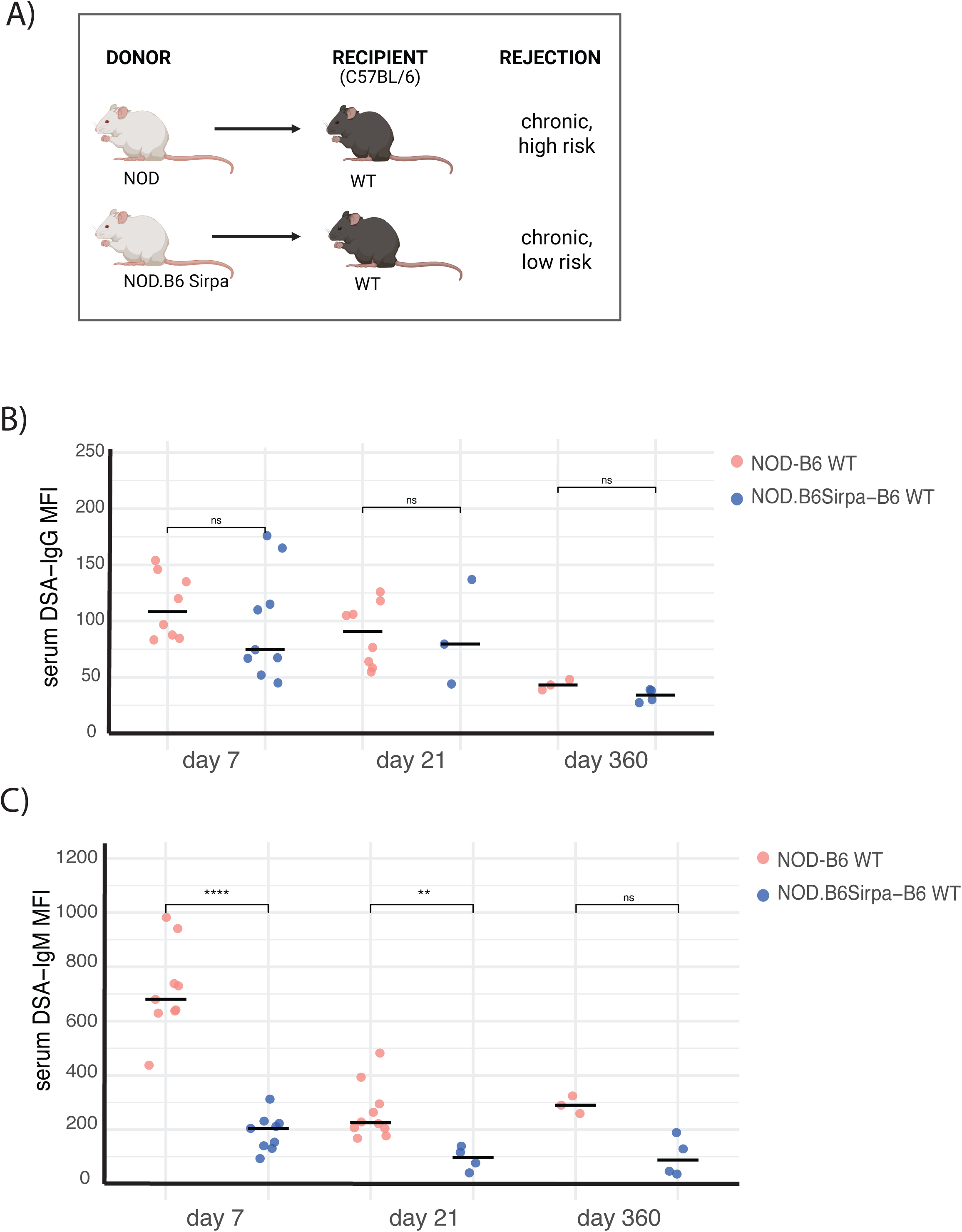
**Serum DSA-IgM but not DSA-IgG is significantly reduced in murine models of lower rejection risk (CD47 ^-/-^ or Sirpa polymorphism)** A) Study design showing kidney transplant from donor NOD mice and NOD.C57BL/6 Sirpa to recipient C57BL/6 mice with 2 different genetic backgrounds –WT and CD47^-/-^. B) Serum DSA-IgG MFI values in NOD-B6 WT and NOD.B6 Sirpa-B6 WT and NOD-B6 CD47^-/-^ at day 7, day 21 and day 360. ns= non-significant. In red are values from NOD-B6 WT mice and in blue are values from NOD.B6 Sirpa-B6 WT mice. Significance was tested by Mann-Whitney *U* test. C) Serum DSA-IgM MFI values in NOD-B6 WT and NOD.B6 Sirpa-B6 WT and NOD-B6 CD47^-/-^ at day 7, day 21 and day 360. ns= non-significant. In red are values from NOD-B6 WT mice and in blue are values from NOD.B6 Sirpa-B6 WT mice. ** = p-value <0.01, **** = p-value < 0.0001, ns= non-significant. Significance was tested by Mann-Whitney *U* test.

## Discussion

In this study, we identify key humoral signatures that differentiate DSA+ patients with kidney allograft rejection compared to those without rejection. Recent studies have highlighted phenotypic differences between these groups including disparities in graft survival, treatment response, histopathology, and clinical outcomes. These findings, though compelling, stop short of explaining why only a subset of DSA+ patients progress to AbMR. Our study addresses this critical gap in our understanding by utilizing novel tools to elucidate features of DSA not currently monitored in the clinic, but that may influence distinct outcomes.

We creatively adapt an interpretable machine learning framework to uncover robust, multivariate associations between antigen-specific humoral profiles and rejection outcomes. Although prior studies have linked AbMR with elevated total IgG and certain subclasses (e.g., IgG1, IgG3, IgG4) (*6*, *11*, *13*), as well as glycosylation patterns such as afucosylation on certain isolated antigen-specific DSAs, our work significantly extends this by developing and employing a highly multiplexed, pan-antigen antibody Fab + Fc coupled profiling strategy. This allows us to generate comprehensive biophysical profiles of DSAs and use them with more nuanced computational approaches to move beyond traditional biomarker prediction and toward identifying putative causal mechanisms driving AbMR, across both early and late rejection stages.

Notably, our study is the first to systematically reveal a significant association between elevated class I-specific IgM responses in AbMR among DSA+ individuals. The uncovered role of IgM Abs in AbMR, especially class-I specific responses at the late time point, is novel. The strength of our approach lies in combining deep humoral profiling with interpretable machine learning to extract mechanistically relevant antibody features and validating these findings across geographically distinct patient cohorts. In doing so, we provide novel insights into the functional antibody landscape of AbMR and advance a generalizable framework for dissecting complex immune-mediated rejection processes.

Our results reveal a striking transition in the humoral landscape from early to late AbMR. In the Pitt cohort, early rejection was likely a result of the memory response against the allograft. Early rejection is characterized by a polyfunctional, class-switched response involving IgM, IgG1, and IgG3, whereas late rejection shifts toward an IgM- and C1q-centric profile. There is evidence suggesting that graft injury and loss result from a cascade of antibody-dependent effector functions, including complement deposition (ADCD) (*30–33*) and cellular cytotoxicity (ADCC) (34–41). In line with these mechanisms, our study recapitulates key molecular drivers of early rejection, as evidenced by elevated IgM, IgG3, and IgG1 profiles (**Fig. 2C**). Importantly, we show that elevated IgM is not merely a secondary marker but plays an active role alongside IgG subclasses in early AbMR among DSA+ patients. This aligns with prior work (*16*, *42*), which speculated that IgM allo-antibodies may serve as early indicators of alloimmunity, potentially preceding the development of de novo IgG DSAs. Functionally, IgM is the most potent initiator of complement, followed by IgG3 and IgG1 (*43*). Beyond complement activation, IgG3 can engage FcγR3A on NK cells and FcγRI/IIA on phagocytes, promoting endothelial damage through ADCC and phagocytic pathways. These mechanistic insights are reinforced by the positive correlation between Z_E_1, Z_E_2 and histological activity scores (**Fig. 2E**), suggesting that these capture antibody features associated with acute microvascular inflammation and complement-mediated endothelial injury.

The humoral landscape in late AbMR reveals an even more skewed profile, with a striking predominance of elevated IgM in DSA+ patients compared to early rejection. While IgM is a potent activator of the complement cascade, its role in graft rejection has remained controversial, contributing to a longstanding diagnostic blind spot. Early observations suggested a protective role: an older study reported that IgM allo-antibodies targeting donor lymphocytes could support long-term graft survival (*17*).However, subsequent findings have been mixed. For instance, Babu et al. (*15*) found that post-transplant IgM DSAs were not associated with rejection per se, but were linked to increased risk of graft failure. In contrast, Everly et al. (*44*) concluded that only IgG DSAs—not IgM DSAs—were associated with poor graft survival. Notably, these earlier studies did not stratify patients by time from transplant. Our results add a new layer to this debate by showing that IgM antibodies are elevated in both early and late AbMR, with particularly strong enrichment in late rejection. Moreover, we observe a moderate positive correlation between our IgM-dominant latent factor (Z_L_1) and histological activity scores, which quantify the extent of graft injury at the time of biopsy.

We also report a strong correlation between an IgG-dominant factor (Z_L_4) and chronicity scores, consistent with the role of IgG in driving structural damage to the graft over time. Since chronicity scores have been significantly associated with graft loss (*45*), our findings support and extend those of Everly et al., indicating an association of IgG in chronic rejection. Yet, our data also suggest that IgM may play a more central pathogenic role in late, rather than early, AbMR. Additional evidence supporting this hypothesis includes lower serum C1q levels in late rejectors— possibly due to consumption and depletion through persistent IgM-mediated complement activation in the allograft —and higher C4d deposition compared to the non-rejectors (late post-transplant) (fig.S7.A), a hallmark of classical complement pathway activation. Moreover, in a murine model of chronic rejection, we observe a significantly greater titer difference for serum IgM-DSAs than IgG-DSAs between high-risk and low-risk mice (**Fig. 5A-C**), further suggesting a potential role of IgM in late post-transplant rejection. Collectively, these findings indicate a shift from IgG-driven Fc-mediated innate responses in early rejection to IgM-mediated complement-dominant mechanisms in late AbMR. Our results thus motivate the need for deeper tissue profiling (capturing proteomic, transcriptomic and spatial information) of allografts to uncover these complex mechanisms underlying AbMR.

A recent study using a semi-supervised clustering algorithm, developed a tool to evaluate disease activity in chronic pathology of kidney allografts (*46*). While the data-driven algorithm was based on Banff lesion scores, a follow-up study revealed uncertainties associated with the predicted outcomes (*47*). Inspired by the need to formulate a clinically potent measure that combined physiologically relevant signatures of chronicity while also capturing underlying mechanistic relationships, we built on the multivariate signatures of elevated IgM and IgG sialylation to compute a combined risk score. This score could stratify patients at risk for late AbMR. The integrative score demonstrated robust classification performance (AUROC = 0.861, **Fig. 3H**), with high sensitivity maintained even at stringent specificity thresholds. This suggests its potential as a clinically actionable tool for early stratification of patients at risk for late AbMR, minimizing false positives while maximizing detection accuracy. Our results thus suggest that routine clinical monitoring of the IgM and sialylation humoral signatures alone, in patients with DSA, could be an effective strategy in early detection of late AbMR (**Fig. 7**).

**Figure 7.**
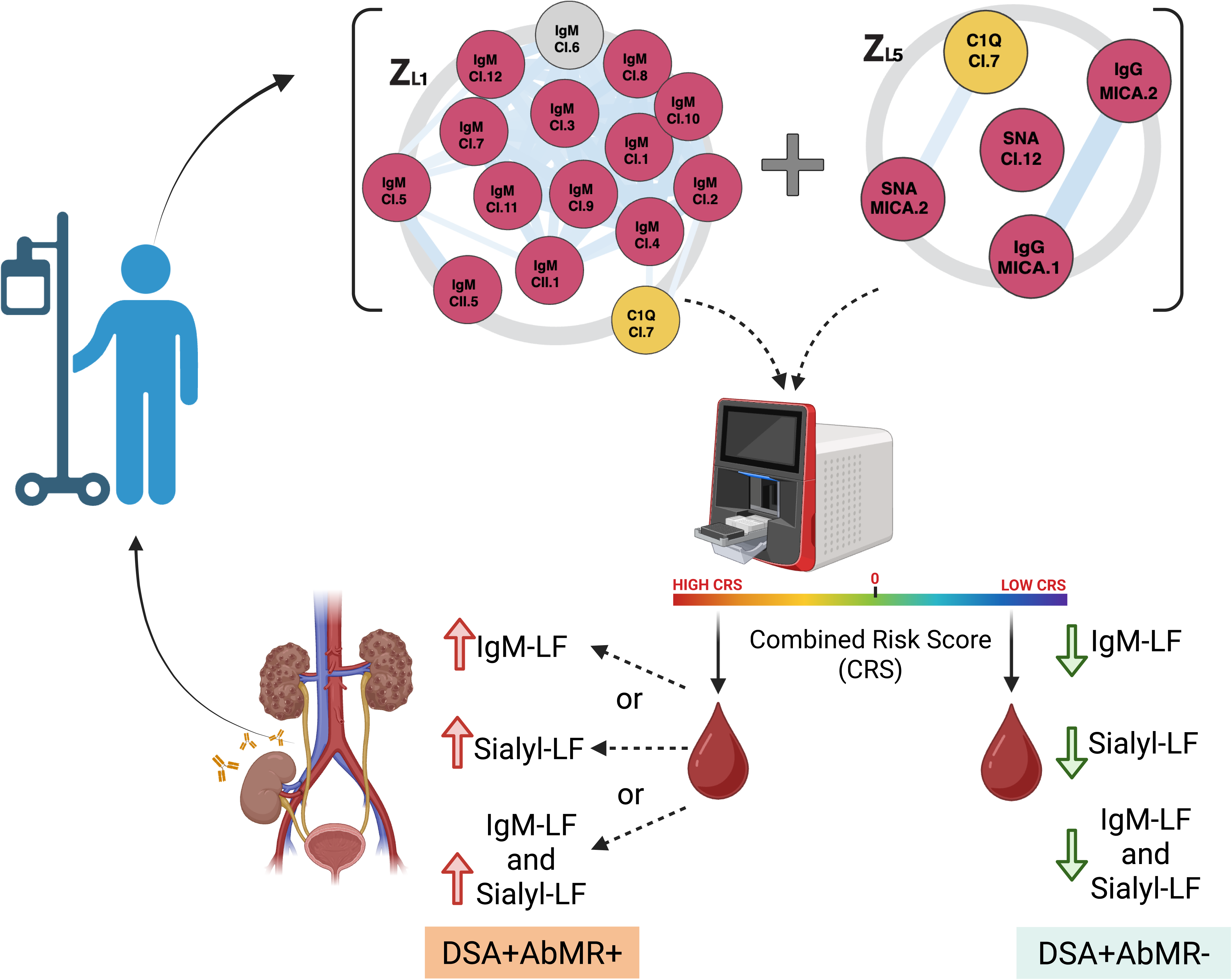
Schematic depicting clinical translation of novel IgM and sialylation signatures in late AbMR risk diagnosis. Workflow showing clinical implementation of the combined risk score for clinical diagnosis of late post-transplant AbMR owing to elevated IgM-LF or elevated sialylation-LF or both.

To better understand the role of IgM in AbMR, several studies have examined its cellular origins, identifying two main B cell sources: B1 and B2 lineages. B1 cells, which are innate-like, are the predominant producers of natural IgM. These antibodies are typically low-affinity, broadly reactive, and serve as part of the early immune response (*48*). However, B1 cells have been shown to play a dual role depending on context, including both protective and pathogenic functions. A recent single-cell transcriptomic study of kidney allograft-infiltrating B cells revealed a B1-like signature associated with local tissue destruction (*49*). Another study demonstrated that the IL-10/TNF-α ratio secreted by transitional B1 cells could serve as a sensitive biomarker for kidney graft rejection (*50*). In lung transplantation, a triple-positive B cell subset (Bhlhe41⁺ Cxcr3⁺ Itgb1⁺), expressing classical B1 markers, was identified as the precursor of *Mzb1*⁺ plasma cells implicated in bronchiolitis obliterans syndrome and poor graft outcomes (*51*). Supporting these findings, our post-hoc analysis reveals strong positive correlations between the IgM-dominant latent factor Z_L_1 and cytokines such as IL-10 and CXCL10—a ligand of Cxcr3—suggestive of a possible B1-driven mechanism in late AbMR (fig.S7.C). However, further studies inspecting transcriptomic profiles of late AbMR allograft tissues would be required to compare the role of B1 versus B2 cells.

Shifting focus to DSA specificities underlying early AbMR, we observed that elevated IgG responses were notably enriched against donor-specific class II HLA antigens, including DR, DQ, and DP serotypes. This was unexpected based on prior studies having predominantly associated class I HLA with early AbMR (*2*, *52*). Even more striking, in late AbMR, we identified a clear enrichment of class I HLA-specific DSAs—particularly within the elevated IgM profiles—despite abundant reports linking late rejection primarily to class II DSAs, especially against DQ and DR. Interestingly, our findings align with an earlier study (*17*), which reported a strong correlation between IgM and class I HLA in the context of AbMR. Our results expand upon this by showing that although class II DSAs remain associated with late AbMR, their enrichment is restricted to the total IgG compartment. In contrast, class I DSAs dominate the IgM profile during late rejection. Moreover, we demonstrate that class II DSA enrichment in the IgG fraction is not limited to the traditionally implicated DQ and DR serotypes but also includes DP serotypes—an observation that has been largely overlooked in previous literature, likely due to assay sensitivity limitations. These novel insights underscore the depth and resolution of our multiplexed profiling approach and highlight the need for renewed investigation into the role of anti-class I IgM in the pathogenesis of late AbMR.

It is well established that beyond class switching, antibody function is further diversified through glycosylation modifications. However, due to the technical complexity, high sample requirement, high cost of traditional glycosylation assays for antigen-specific antibodies, and resultant limited data availability, the role of antibody glycosylation in AbMR has remained relatively obscure until recent years. Emerging studies have begun to shed light on this layer of immune regulation. For example, Bharadwaj et al. (*11*) isolated anti-HLA-A2 IgG antibodies and demonstrated that elevated afucosylation was associated with enhanced FcγR3A-mediated cytotoxicity. In contrast, Barba et al. (*19*) reported no significant association between sialylation and AbMR. Challenging this, Pernin et al. (*13*) found that low sialylation of de novo IgG3 antibodies was linked to a higher risk of AbMR. Beyond AbMR, higher sialylation has also been linked to increased breadth and higher affinity of vaccine-mediated antibody responses (*53*). Overall, these studies suggest a more nuanced role for glycosylation in graft rejection and underscore the need for development and adoption of more comprehensive pan-antigen glycoprofiling techniques.

In our study, without relying on prior biological assumptions, harnessing our sample-sparing, highly multiplexed lectin-based antigen-specific pan-Ig antibody glycoprofiling method (*54*), we identify a potentially pro-inflammatory role for sialylation in both early and late AbMR. While sialylation is canonically viewed as anti-inflammatory, its role in complement activation and AbMR remains debated. Our multivariate analysis reveals that elevated sialylation is moderately correlated with both activity and chronicity scores in early AbMR—suggesting a contribution to ongoing immune injury and structural damage. This role becomes more pronounced in late AbMR, where sialylation and total IgG levels show strong associations with chronicity and moderate associations with activity, indicating their potential involvement in long-term tissue remodeling and damage. Intriguingly, our comparison of early and late rejection time points uncovers a distinct shift: galactosylation, particularly in class I antigen-specific antibodies, becomes more prominent in late rejection. Galactosylation has earlier been shown to promote complement activation via hexamerization of IgG1 (*55*). To our knowledge, this is the first study to directly compare galactosylation patterns across rejection stages. It is worth highlighting here again in this context that our lectin-based glycoprofiling technique does not depend on isolation of specific antibody isotypes nor antigen-specificities and thus provides the broadest readout of antibody glycoprofiles. Our findings thus underscore the need for future targeted MS-based analyses of glycosylated class I DSAs, along with functional assays, to dissect the mechanistic role of galactosylation in the pathogenesis of early versus late AbMR.

In conclusion, this study resolves long-standing contradictions regarding the association of key humoral signatures with both early and late AbMR by employing a comprehensive antibody Fab + Fc profiling method and an unbiased and statistically rigorous computational approach. Moving beyond conventional biomarker prediction, we uncover putative causal signatures that not only reveal novel mechanistic insights but also address diagnostic blind spots across stages of AbMR. Validation in geographically distinct patient cohorts further underscores the robustness of these signatures, which—when monitored early post-transplant—can predict later rejection events (**Fig. 7**). Our findings thus advocate for deeper investigations into the cellular and spatial-transcriptomic dynamics underpinning graft rejection. Importantly, while focused on kidney transplantation, the methodological framework we present is broadly applicable to other transplant and immune-mediated conditions.

## Methods

### Study design

The discovery cohort (Pittsburgh cohort 1) included samples from patients who underwent kidney transplantation between January 2013 and December 2017 at University of Pittsburgh Medical Center and who were recruited to participate in a biorepository initiative at Thomas E. Starzl Transplantation Institute (STI). All patients signed a written informed consent (institutional review board numbers STUDY19100380). A total of 530 patients were screened for the following immunologic events: presence of post-transplant DSA and biopsy-proven ABMR. For this study we identified a total of 36 patients who developed DSA in the first 24 months post-transplant defining two study groups: patients with ABMR (DSA+AbMR+, n=19) and patients without ABMR (DSA+AbMR−, n=17). The first group (DSA+AbMR+) comprised patients who developed AbMR post-transplant including 15 who had TCMR as well, and DSA+AbMR-group with 1 patient having TCMR Besides patients with no rejection, those who had borderline rejection (but not AbMR) were included in the DSA+AbMR-group. AbMR was detected by kidney allograft protocol and indication biopsies. Clinical data of the study patients were extracted from the prospective database of the STI. Details of other diagnoses and treatment regimen followed for patients in this cohort can be found in Supplementary Table 1 and 2. In addition, patients were followed until May 1, 2019, and allograft loss was assessed. This cohort of patients was further stratified into early (n(DSA+AbMR-) = 8, n(DSA+AbMR+) = 10) and late events (n(DSA+AbMR-) = 9, n(DSA+AbMR+) = 9) based on the threshold of 6 weeks from time of transplant, for further analyses.

For validating higher C1q consumption (by proxy of higher tissue C4d deposition), we used a second group of samples from the same biorepository. This included a total of 81 patients categorised as DSA+AbMR+ (n=31) and DSA+AbMR-(n=50) patients based on the 6 week post time from transplant cut-off to match those in the late group within the discovery cohort.

### Validation cohort

The validation cohort (Toronto cohort) included 32 samples, including 16 DSA+AbMR+ and 16 DSA+AbMR-. Using the CoReTRIS registry, kidney transplant recipients from the period between 2007 and 2022 were selected. Serum samples were collected post-transplant, at the time of a for cause kidney allograft biopsy, and donor-specific anti-HLA antibodies were assessed using a Luminex single-antigen bead assay as part of standard clinical care. For each patient, for-cause 18-gauge needle biopsies were collected and scored by a renal pathologist according to the Banff 2022 classification criteria. Inclusion criteria for this study were patients who had at least one DSA against their current kidney graft at the time of biopsy and who had a diagnosis of acute active AMR, chronic active/inactive AMR, mixed AMR/cellular rejection, or no rejection based on the Banff criteria. Exclusion criteria were patients who were negative for DSA or who had pure cellular rejection. This study was approved by the University Health Network institutional research ethics board (Coordinated Approval Process for Clinical Research [CAPCR] identifier 18-5489). Clinical metadata is as shown in Supplementary table 3.

### Sample preparation

Serum samples were stored at –80°C until analysis. Prior to use, samples were thawed on ice and prepared according to the intended probe type. For the multiplexed antibody-binding assays (e.g., total IgG, IgG1, IgG3, IgM, FcR2B, FcR3A, and lectin-based glycosylation probes such as SNA and RCA), samples were transferred to 96-well U-bottom plates and diluted in assay buffer (1× PBS, 0.1% BSA). A 1:50 dilution was used for IgG4 and lectin-binding measurements, while a 1:250 dilution was used for all other probes. These dilutions were determined based on preliminary titration curves using pooled serum samples to ensure optimal signal-to-noise ratio and assay performance. 1× PBS buffer was used as a technical negative control. Further for positive and negative controls, positive and negative control beads provided by One Lambda were also used as described below. For the C1q complement-binding assay, serum was not diluted and underwent heat inactivation at 56°C for 30 minutes prior to use, to eliminate endogenous C1q complexes. Following this, recombinant C1q protein was diluted in HEPES buffer and mixed in equal volume with the heat-inactivated serum. This serum–C1q mix was incubated with 5 μL of LABScreen antigen-coated beads for 20 minutes. Without washing, 5 μL of anti-C1q-PE probe was then added and incubated for an additional 20 minutes. Afterward, 80 μL of PBS was added to each well, followed by a single centrifugation and flicking step. No intermediate wash steps were performed prior to reading. All samples were acquired on a Cytek Aurora flow cytometer, and data were analyzed using FlowJo software.

### One Lambda Beads and Multiplexed Assay

LABScreen Mixed HLA Class I, Class II, and MICA pre-coated beads were purchased from One Lambda. The mixed beads included 21 specific bead regions with the following subdivisions: 1 positive control, 1 negative control, 12 class I-specific antigens, 5 class II-specific antigens, and 2 MICA-specific antigens. Each bead ID is identified by a cluster of specific HLAs and emit a specific red wavelength. Bead analysis, including gating and MFI determination, was conducted in FlowJo software. The multiplexed assay was performed as per the manufacturer’s recommendations and as previously reported by Brown et al, 2012(*53*) and lectin-binding assays were performed as reported by us earlier(*8*, *54*). Briefly, the LABScreen beads were vortexed before adding 1.3uL per sample into the assay buffer (1X PBS, 0.1%BSA) to make a working solution with an approximate concentration of 50 beads/well. Next, the beads were incubated with diluted serum for 1hr. After incubation, 1X LABScreen wash buffer was added to each well and centrifuged to pellet the beads before removing the wash buffer by flicking. Next, the beads were incubated with a fluorescently labelled probe for 30 minutes. For this study, the following human-specific probes were measured: total IgG, IgG1, IgG3, IgM, FcR2B, FcR3A, SNA (sialic acid binder), RCA (galactose binder), and C1q. For isotype, subclass, and FcR analysis, the probe was diluted in the assay buffer. For glycosylation analysis, the lectins were diluted in lectin buffer (1X PBS, 20mM Tris, 0.1mM Ca, 0.1mM Mg, 0.1mM Mn, 0.1% BSA). After probe incubation, the plate is washed (3x) and resuspended in 1X PBS before analysed on a flow cytometer (Cytek Aurora).

### Complement Analysis

The One Lambda C1qScreen assay was used to identify the presence of complement-binding (C1q) antibodies and performed according to the manufacturer’s instructions. Undiluted patient serum was heat inactivated to break up the C1q complex already present. Next, the total volume of C1q was calculated (1ul C1q/well) and diluted in HEPES buffer. Equal parts of undiluted serum and diluted C1q were mixed together before adding the combined solution to 5ul of antigen-coated microbeads for 20 minutes. Without washing, 5uL of anti-C1q-PE was incubated with the beads for 20 minutes. Next, 80uL of PBS was added to each well and centrifuged before flicking. Finally, the fluorescence was measured on the Cytek Aurora.

### Cytokine profiling

Cytokine quantification of the serum samples was performed using a 12-plex Luminex Discovery Assay (Biotechne LXSAHM-12). The cytokines were as follows: TNF-α, IL-6, IL-8/CXCL8, CXCL10/IP-10, IL-10, CCL13/MCP-4, TSLP, CXCL9/MIG, TIE-2, ICAM-1/CD54, HGF, and CXCL1. Serum samples were diluted by a factor of 2 and the assay was performed according to the manufacturer’s protocol. Cytokines with median average MFI over 13 were used for further analysis.

### SLIDE analysis

We analysed Ab profiles against antigen minipools (fig.S1) using SLIDE. We filtered out features with a median MFI of < 50, to remove features below the limit of detection. The outcomes were assigned a binary label (DSA+AbMR+= 1, DSA+AbMR-= 0) for both early and late sub-groups. FcR2B was not included for the multivariate analyses. The SLIDE model involved tuning the two hyperparameters: delta and lambda, where delta determines the total number of latent factors (*k*) and lambda determines the composition of these latent factors. Delta and lambda tuning was performed using a grid search as previously described(*18*). To prevent overfitting and confirm the robustness of the latent factors identified, our analytical pipeline incorporated multiple layers of statistical control. Initially, dimensionality reduction was carried out through ER to extract latent factors (Zs) from the original feature space (Xs), with this transformation backed by formal identifiability assurances. These latent factors were then used in a supervised regression framework, where false discovery rate (FDR) was tightly controlled, and all hypothesis testing adhered to a significance level of α = 0.05 FDR tightly controlled at a threshold of 0.1. Additionally, to further validate model generalizability and prevent overfitting, we employed a nested k-fold cross-validation strategy coupled with permutation testing. Model significance was assessed using permutation testing (in a matched k-fold cross-validation framework as previously described, as well as against a null model with randomly selected latent factors. For cases where interactions were significant in the model, the permutation test was performed against a null model of randomly selected interactions with other latent factors. Only the model for distinguishing late rejectors from late non-rejectors, included significant interactions between latent factors.

The significant latent factors were further pruned post-hoc to retain only those features with high loadings. Only those latent factors that had a significant effect size (absolute cliff’s delta > 0.12) were used for post-hoc analysis and interpretation. These latent factors were visualized as correlation networks based on corresponding features in these factors (identified from the allocation matrix). These networks were visualized using the qgraph package in R.

### SLIDE cross-prediction

SLIDE factorizes the feature matrix from the discovery cohort (denoted as X_D_, with dimensions n₁ × p, where n₁ is the number of samples and p is the number of humoral features) into two components: a latent factor matrix Z_D_ (n₁ × k, representing k latent dimensions, where k ≪ p), and an allocation matrix A_D_ with dimensions k × p as in Eq.1

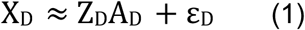

Here, ε represents the random error term. SLIDE then used a knockoff-based framework to identify latent factors from the discovery cohort that were significantly associated with the outcome label. These selected latent factors were subsequently regressed onto the outcome within the discovery cohort to assess their predictive relevance (Eq.2)

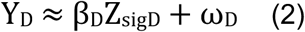

Z_sigD_ = {feature selected by knockoff}, and ω represents random error. Using only the significant latent factors for outcome prediction (Y) and retaining the allocation matrix (A_D_) learned from the discovery cohort, we reconstructed the feature matrix in the validation cohort as described in Eq.3.

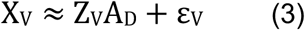

Retaining the model trained on the discovery cohort, we assessed disease outcomes in the validation cohort (X_V_, with dimensions n₂ × p, where n₂ is the number of validation samples and p is the number of features), as outlined in Eq.4.

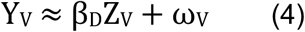

Interaction terms were also used in the regression equation only in the model differentiating outcome labels in the late subgroup, as the model performed best with the interaction terms included. ROC curves were generated using the **pROC** package in R, based on predicted and true labels from the validation cohort. For comparison, we also evaluated model performance in the discovery cohort by computing AUROCs using the same model, applied to its corresponding predicted and actual labels.

### Latent factor correlation with Activity and Chronicity scores

Activity and chronicity indices were calculated using Banff scores based on the definition previously established(*24*) where, Activity index = Banff glomerulitis (g) + peritubular capillaritis (ptc) + arteritis (v) + C4d scores, with a maximum score of 12; Chronicity index = 2 x (interstitial fibrosis (ci) + tubular atrophy (ct) + chronic vasculopathy (cv) + chronic glomerulopathy (cg)), with a maximum score of 12.

The indices were calculated for the Toronto cohort samples only, based on data availability. The spearman’s rank correlation was calculated between each index and the significant latent factors. The statistical significance of these correlations was determined using permutation-derived P values, which were subsequently adjusted for multiple testing using the Benjamini-Hochberg procedure.

### Relationship between cytokines and significant latent factors from SLIDE

Spearman’s rank correlation was used to calculate correlation coefficient between cytokines that passed the filtering criteria. The statistical significance of these correlations was determined using permutation-derived P values, which were subsequently adjusted for multiple testing using the Benjamini-Hochberg procedure.

### Combined risk score

Latent factor values for each sample in the validation cohort were computed by fitting the model learned from the discovery cohort (as described in cross-prediction). The combined risk score for patient i was defined as in Eq. 5;

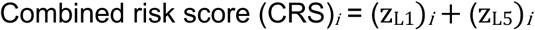

### Probe-antigen enrichment

Historical DSA specificities developed by patients from the time from kidney allograft transplant till sampling date was recorded. Antigens to which DSAs were developed in AbMR+ patients were noted as rejector antigens, and DSAs to which alloantibodies were developed in AbMR-patients (and not in AbMR+ patients) were noted as non-rejector antigens. Those specificities that showed up in case of both AbMR+ patients as well as AbMR-patients were retained in the list of rejector antigens (and dropped from the list of non-rejector antigens) to reduce any false negatives. MICA specificities were not considered for this analysis. All specificities were considered independently and separately for early and late subgroups. The significance of enrichment of humoral signatures (for early/late) in anti-HLA specificities (class I and II) was calculated using hypergeometric test, for every probe (antibody isotype/subclass, FcR, lectin). Membership of an antigen in a minipool was recorded as a unit count, for antigens present on more than 1 minipool. The threshold for significance was kept at P<=0.05. Further, only those probe-HLA pairs were retained where the antigen had membership in more than 2 minipools. HLA specificities that showed a significant enrichment but were not present in any donor were designated as non-DSA anti-HLA.

### Reagents

FITC-labeled anti-mouse IgG and rhodamine red–conjugated anti-mouse IgM were used for measuring the serum IgM and IgG DSA in mice.

### Mice

C57BL/6J (B6 WT) and NOD/ShiLtJ (NOD) mice were purchased from The Jackson Laboratory. NOD.NOR-Ila-D2Gul482 (NOD.B6 Sirpa) mice were generated and provided by Jayne S. Danska (University of Toronto).

### Transplant Surgery

NOD or NOD.B6 Sirpa mice were used as kidney donors and B6 WT mice were used as recipients. Recipients underwent bilateral nephrectomy at the time of transplantation.

### Experimental Groups

B6 WT mice received kidneys from either NOD donor mice (group NOD-B6 WT) or NOD.B6 Sirpa donor mice (group NOD.B6 Sirpa-B6 WT).

### Circulating donor-specific antibodies assay

The levels of circulating anti-donor DSAs of IgG and IgM in recipient sera at indicated time points were assessed by flow cytometry as described previously (add citation). Briefly, recipient sera were incubated with C3H donor splenocytes at 37℃ for 30 minutes, and washed cells were then incubated with FITC-labeled anti-mouse IgG and rhodamine red–conjugated anti-mouse IgM at 4℃ for 1 hour. Cells were analyzed by flow cytometry with results expressed as mean fluorescence intensity to reflect individual serum DSA levels.

## Data Availability

All data produced in the present study are available upon reasonable request to the authors

## Code Availability

Code for all analyses and figure generation can be found at https://github.com/TrirupaChakraborty/ABMR.git

**Supplementary Figure 1** A) HLA-class I and class II antigen to minipool IDs. Minipools in yellow correspond to HLA class I pools and those in blue correspond to HLA class II pools. (B-F) Differences in clinical parameters between DSA+AbMR-and DSA+AbMR+ patients.

**Supplementary Figure 2** A) Violin-plot showing log (MFI) values of total IgG titers against all antigen pools (minipools) used in the study. (MFI = mean fluorescent intensity). CI = indicates a minipool with class I antigens, CII = indicates a minipool with class II antigens. Refer to antigen minipool representation and mapping to antigens in supplementary material in (A). ns = non-significant B) Performance of the real SLIDE model (in purple; AUROC =0.55) in discriminating between DSA+AbMR+ and DSA+AbMR-patients in the Pittsburgh cohort, relative to the distribution of the performance of models built using permuted (shuffled) outcome labels (in green). Model performance was measured in a *k*-fold cross-validation framework with permutation testing, and the metric used is AUROC. ns = not significant using the Mann-Whitney *U* test.

**Supplementary Figure 3** Univariate MFI values specific to antibody features captured by significant LFs in the early group of patients from the Pittsburgh cohort (A-I), across antigen minipools.

**Supplementary Figure 4** Univariate MFI values specific to antibody features captured by significant LFs in the early group of patients from the Toronto cohort (A-I), across antigen minipools.

**Supplementary Figure 5** Univariate MFI values specific to antibody features captured by significant LFs in late group of patients from the Pittsburgh cohort (A-I), across antigen minipools.

**Supplementary Figure 6** Univariate MFI values specific to antibody features captured by significant LFs in late group of patients from the Toronto cohort (A-I), across antigen minipools.

**Supplementary Figure 7.** A) Violin plots showing C4d scores in patients beyond 6 weeks post transplant, from pittsburgh cohort 2. Red violin corresponds to the DSA+AbMR+ patients and the yellow violin corresponds to DSA+AbMR-patients. Plot points in red signify a C4d score of 2 or more. **** indicates *P* < 0.0001 using the Mann-Whitney *U* test. B) Scatter plot with IgM LF scores on the y-axis and sialylation LF scores on the x-axis. Samples from Pitt and Toronto cohort are plotted on the same graph, with circles corresponding to the Pitt samples (Discovery cohort) and triangles corresponding to the Toronto samples (Validation cohort). Red denotes samples from DSA+AbMR+ patients and yellow denotes samples belonging to DSA+AbMR-patients. C) Heatmap showing spearman’s correlation between late latent factors and cytokine (average MFI) pass the *P* < 0.05 threshold. Color gradient on the right maps blue to correlation coefficient of -0.6 and red to +0.6. Correlations in gray were not significant.

**Supplementary Figure 8.** Probe-minipool features selected by significant latent factors, enriched in patient DSAs (serotypes). Features that were elevated in DSA+AbMR+ (rejector) patients as compared to the DSA+AbMR-(non-rejector) patients in the latent factors from early (A) and late groups (B) respectively, are depicted in red in the SLIDE column, and those that were lower in early non-rejectors is depicted in yellow. Patient DSA serotypes that are in red are enriched in the rejectors for the corresponding probe (in the probe-minipool pair). Patient DSA serotypes that are in yellow are enriched in the non-rejectors for the corresponding probe (in the probe-minipool pair). Patient serotypes in blue boxes are non-DSA anti-HLA enriched in corresponding probe-minipool pairs.

**Supplementary Table 1.** Pittsburgh patient cohort 1 characteristics and relevant metadata.

**Supplementary Table 2.** Pittsburgh patient cohort 1 metadata regarding treatment regimen between early and late groups.

**Supplementary Table 3.** Toronto cohort patient characteristics and relevant metadata.

