## supplementary figures for "High-resolution multiplexed antibody-omics and interpretable machine learning unveil novel pathogenic mechanisms in kidney transplant rejection"

fig.S1)

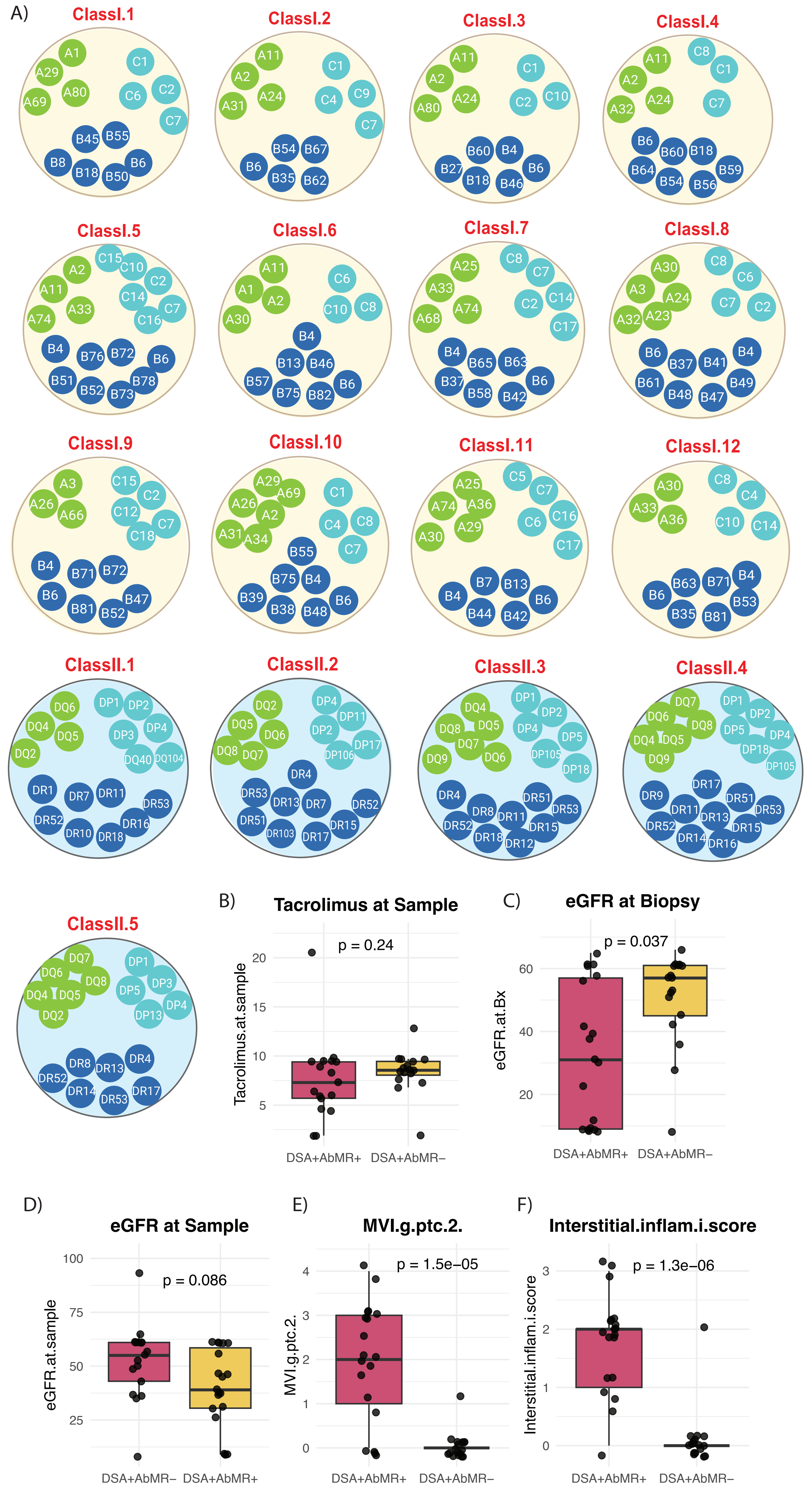
A)
ClassI.1
A1 A29 A80 A69 C1 C6 C2 C7 B45 B55 B8 B18 B50 B6
ClassI.2
A11 A2 A24 A31 C1 C4 C9 C7 B54 B67 B6 B35 B62
ClassI.3
A11 A2 A24 A80 C1 C2 C10 B60 B4 B27 B18 B46 B6
ClassI.4
A11 A2 A24 A32 C8 C1 C7 B6 B60 B18 B64 B54 B56 B59
ClassI.5
A2 A11 A33 A74 C15 C10 C2 C14 C7 C16 B4 B76 B72 B6 B51 B52 B73 B78
ClassI.6
A11 A1 A2 A30 C6 C10 C8 B4 B13 B46 B57 B75 B82 B6
ClassI.7
A25 A33 A74 A68 C8 C7 C2 C14 C17 B4 B65 B63 B37 B58 B42 B6
ClassI.8
A30 A3 A24 A23 A32 C8 C6 C7 C2 B6 B37 B41 B4 B61 B48 B47 B49
ClassI.9
A3 A26 A66 C15 C2 C12 C7 C18 B4 B71 B72 B6 B81 B52 B47
ClassI.10
A29 A26 A69 A2 A31 A34 C1 C4 C8 C7 B55 B75 B4 B39 B38 B48 B6
ClassI.11
A25 A74 A36 A29 A30 C5 C7 C6 C16 C17 B7 B13 B4 B6 B44 B42
ClassI.12
A30 A33 A36 C8 C4 C10 C14 B4 B63 B71 B6 B53 B35 B81
ClassII.1
DQ6 DQ4 DQ5 DQ2 DP1 DP2 DP3 DP4 DQ40 DQ104 DR1 DR7 DR11 DR53 DR52 DR16 DR10 DR18
ClassII.2
DQ2 DQ5 DQ6 DQ8 DQ7 DP4 DP11 DP2 DP17 DP106 DR4 DR53 DR13 DR7 DR52 DR51 DR15 DR103 DR17
ClassII.3
DQ4 DQ8 DQ5 DQ7 DQ9 DQ6 DP1 DP2 DP4 DP5 DP105 DP18 DR4 DR51 DR8 DR11 DR53 DR52 DR15 DR18 DR12
ClassII.4
DQ7 DQ6 DQ8 DQ4 DQ5 DQ9 DP1 DP2 DP5 DP4 DP18 DP105 DR17 DR9 DR51 DR11 DR13 DR53 DR52 DR15 DR14 DR16
ClassII.5
DQ7 DQ6 DQ8 DQ4 DQ5 DQ2 DP1 DP3 DP5 DP4 DP13 DR8 DR4 DR13 DR52 DR14 DR53 DR17
B)
Tacrolimus at Sample
p = 0.24
Tacrolimus.at.sample
20
15
10
5
DSA+AbMR+
DSA+AbMR−
C)
eGFR at Biopsy
p = 0.037
eGFR.at.Bx
60
40
20
DSA+AbMR+
DSA+AbMR−
D)
eGFR at Sample
p = 0.086
eGFR.at.sample
100
75
50
25
DSA+AbMR−
DSA+AbMR+
E)
MVI.g.ptc.2.
p = 1.5e−05
MVI.g.ptc.2.
4
3
2
1
0
DSA+AbMR+
DSA+AbMR−
F)
Interstitial.inflam.i.score
p = 1.3e−06
Interstitial.inflam.i.score
3
2
1
0
DSA+AbMR+
DSA+AbMR−

fig.S2)

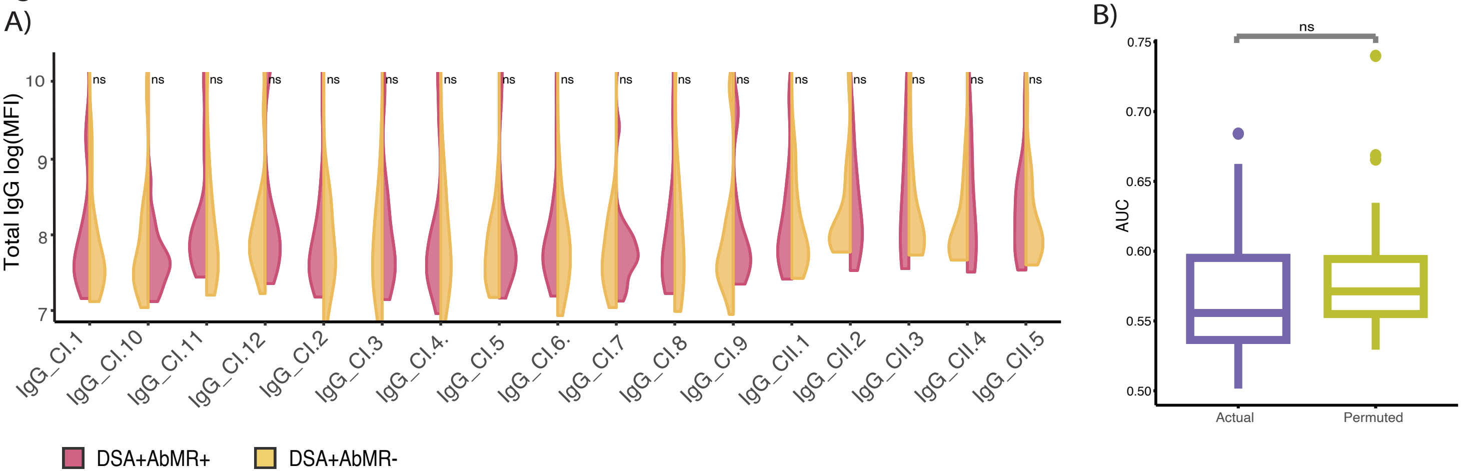
A)
B)
Total IgG log(MFI)
10
9
8
7
ns
IgG_CI.1
IgG_CI.10
IgG_CI.11
IgG_CI.12
IgG_CI.2
IgG_CI.3
IgG_CI.4.
IgG_CI.5
IgG_CI.6.
IgG_CI.7
IgG_CI.8
IgG_CI.9
IgG_CII.1
IgG_CII.2
IgG_CII.3
IgG_CII.4
IgG_CII.5
DSA+AbMR+
DSA+AbMR-
AUC
0.75
0.70
0.65
0.60
0.55
0.50
Actual
Permuted

fig.S3)

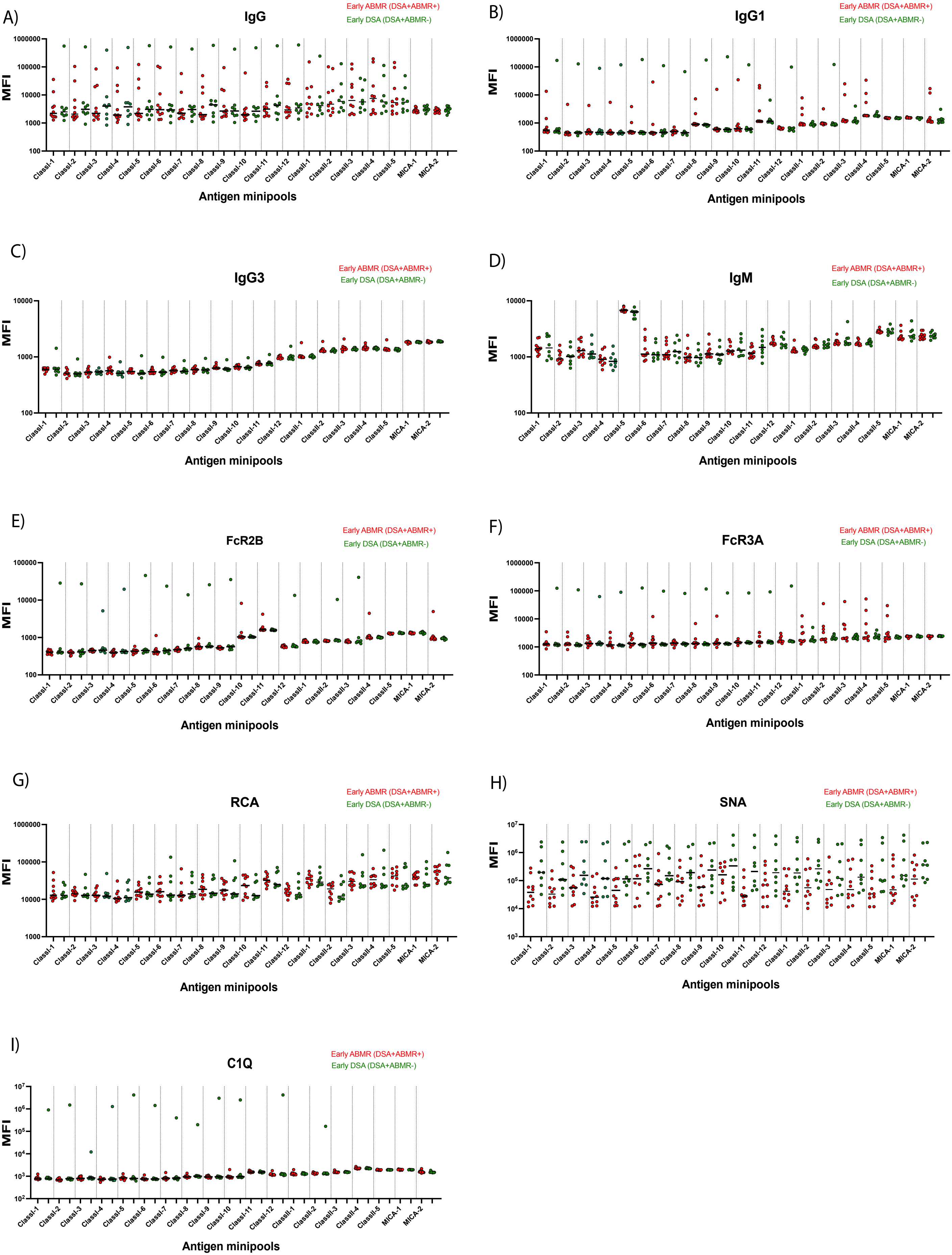

A)
IgG
Early ABMR (DSA+ABMR+)
Early DSA (DSA+ABMR-)
MFI
1000000
100000
10000
1000
100
ClassI-1 ClassI-2 ClassI-3 ClassI-4 ClassI-5 ClassI-6 ClassI-7 ClassI-8 ClassI-9 ClassI-10 ClassI-11 ClassI-12 ClassII-1 ClassII-2 ClassII-3 ClassII-4 ClassII-5 MICA-1 MICA-2
Antigen minipools
B)
IgG1
Early ABMR (DSA+ABMR+)
Early DSA (DSA+ABMR-)
MFI
1000000
100000
10000
1000
100
ClassI-1 ClassI-2 ClassI-3 ClassI-4 ClassI-5 ClassI-6 ClassI-7 ClassI-8 ClassI-9 ClassI-10 ClassI-11 ClassI-12 ClassII-1 ClassII-2 ClassII-3 ClassII-4 ClassII-5 MICA-1 MICA-2
Antigen minipools
C)
IgG3
Early ABMR (DSA+ABMR+)
Early DSA (DSA+ABMR-)
MFI
10000
1000
100
ClassI-1 ClassI-2 ClassI-3 ClassI-4 ClassI-5 ClassI-6 ClassI-7 ClassI-8 ClassI-9 ClassI-10 ClassI-11 ClassI-12 ClassII-1 ClassII-2 ClassII-3 ClassII-4 ClassII-5 MICA-1 MICA-2
Antigen minipools
D)
IgM
Early ABMR (DSA+ABMR+)
Early DSA (DSA+ABMR-)
MFI
10000
1000
100
ClassI-1 ClassI-2 ClassI-3 ClassI-4 ClassI-5 ClassI-6 ClassI-7 ClassI-8 ClassI-9 ClassI-10 ClassI-11 ClassI-12 ClassII-1 ClassII-2 ClassII-3 ClassII-4 ClassII-5 MICA-1 MICA-2
Antigen minipools
E)
FcR2B
Early ABMR (DSA+ABMR+)
Early DSA (DSA+ABMR-)
MFI
100000
10000
1000
100
ClassI-1 ClassI-2 ClassI-3 ClassI-4 ClassI-5 ClassI-6 ClassI-7 ClassI-8 ClassI-9 ClassI-10 ClassI-11 ClassI-12 ClassII-1 ClassII-2 ClassII-3 ClassII-4 ClassII-5 MICA-1 MICA-2
Antigen minipools
F)
FcR3A
Early ABMR (DSA+ABMR+)
Early DSA (DSA+ABMR-)
MFI
1000000
100000
10000
1000
100
ClassI-1 ClassI-2 ClassI-3 ClassI-4 ClassI-5 ClassI-6 ClassI-7 ClassI-8 ClassI-9 ClassI-10 ClassI-11 ClassI-12 ClassII-1 ClassII-2 ClassII-3 ClassII-4 ClassII-5 MICA-1 MICA-2
Antigen minipools
G)
RCA
Early ABMR (DSA+ABMR+)
Early DSA (DSA+ABMR-)
MFI
1000000
100000
10000
1000
ClassI-1 ClassI-2 ClassI-3 ClassI-4 ClassI-5 ClassI-6 ClassI-7 ClassI-8 ClassI-9 ClassI-10 ClassI-11 ClassI-12 ClassII-1 ClassII-2 ClassII-3 ClassII-4 ClassII-5 MICA-1 MICA-2
Antigen minipools
H)
SNA
Early ABMR (DSA+ABMR+)
Early DSA (DSA+ABMR-)
MFI
$10^7$
$10^6$
$10^5$
$10^4$
$10^3$
ClassI-1 ClassI-2 ClassI-3 ClassI-4 ClassI-5 ClassI-6 ClassI-7 ClassI-8 ClassI-9 ClassI-10 ClassI-11 ClassI-12 ClassII-1 ClassII-2 ClassII-3 ClassII-4 ClassII-5 MICA-1 MICA-2
Antigen minipools
I)
C1Q
Early ABMR (DSA+ABMR+)
Early DSA (DSA+ABMR-)
MFI
$10^7$
$10^6$
$10^5$
$10^4$
$10^3$
$10^2$
ClassI-1 ClassI-2 ClassI-3 ClassI-4 ClassI-5 ClassI-6 ClassI-7 ClassI-8 ClassI-9 ClassI-10 ClassI-11 ClassI-12 ClassII-1 ClassII-2 ClassII-3 ClassII-4 ClassII-5 MICA-1 MICA-2
Antigen minipools

fig.S4)

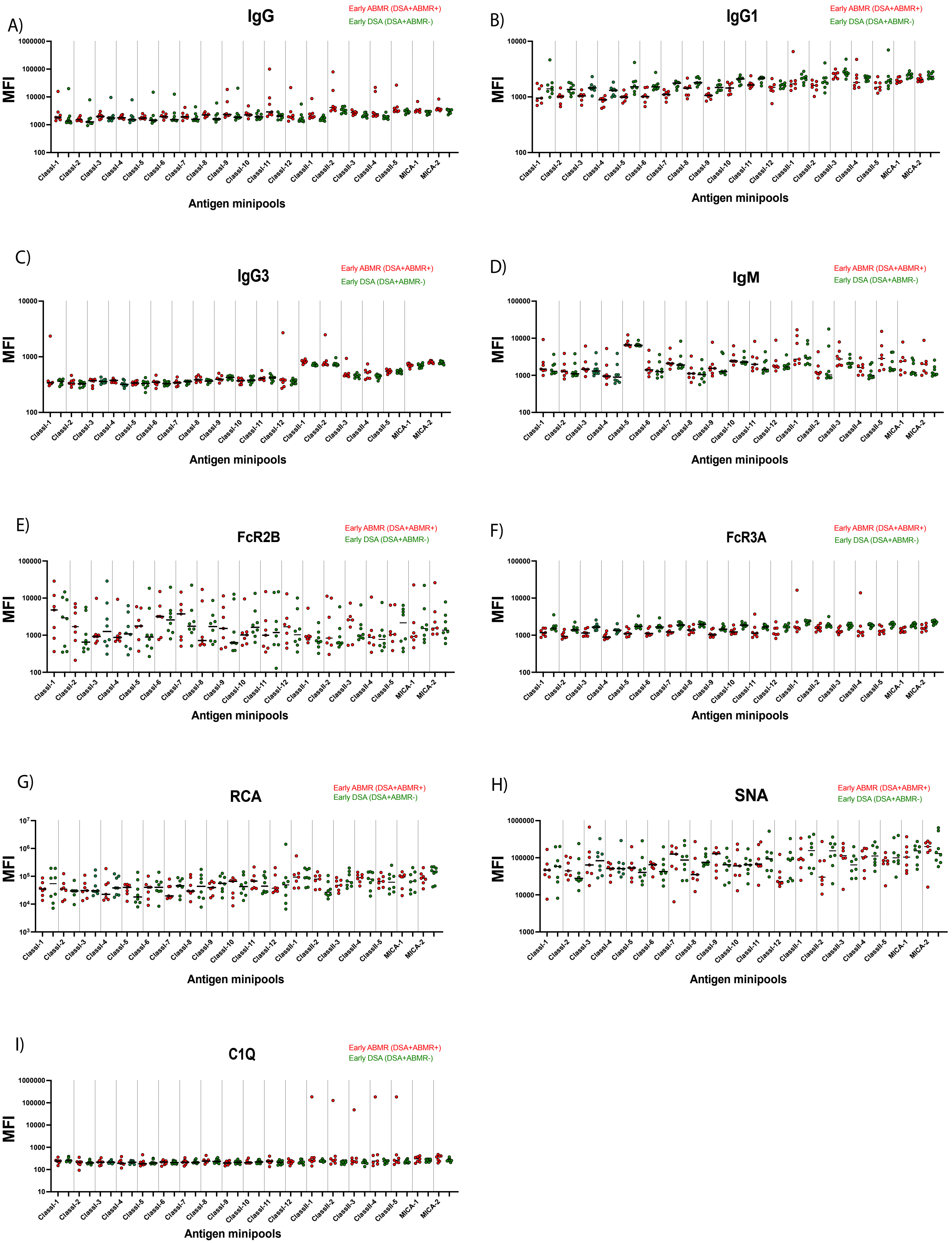
A)
IgG
Early ABMR (DSA+ABMR+)
Early DSA (DSA+ABMR-)
MFI
ClassI-1 ClassI-2 ClassI-3 ClassI-4 ClassI-5 ClassI-6 ClassI-7 ClassI-8 ClassI-9 ClassI-10 ClassI-11 ClassI-12 ClassII-1 ClassII-2 ClassII-3 ClassII-4 ClassII-5 MICA-1 MICA-2
Antigen minipools
B)
IgG1
Early ABMR (DSA+ABMR+)
Early DSA (DSA+ABMR-)
MFI
Antigen minipools
C)
IgG3
Early ABMR (DSA+ABMR+)
Early DSA (DSA+ABMR-)
MFI
Antigen minipools
D)
IgM
Early ABMR (DSA+ABMR+)
Early DSA (DSA+ABMR-)
MFI
Antigen minipools
E)
FcR2B
Early ABMR (DSA+ABMR+)
Early DSA (DSA+ABMR-)
MFI
Antigen minipools
F)
FcR3A
Early ABMR (DSA+ABMR+)
Early DSA (DSA+ABMR-)
MFI
Antigen minipools
G)
RCA
Early ABMR (DSA+ABMR+)
Early DSA (DSA+ABMR-)
MFI
Antigen minipools
H)
SNA
Early ABMR (DSA+ABMR+)
Early DSA (DSA+ABMR-)
MFI
Antigen minipools
I)
C1Q
Early ABMR (DSA+ABMR+)
Early DSA (DSA+ABMR-)
MFI
Antigen minipools

fig.S5)

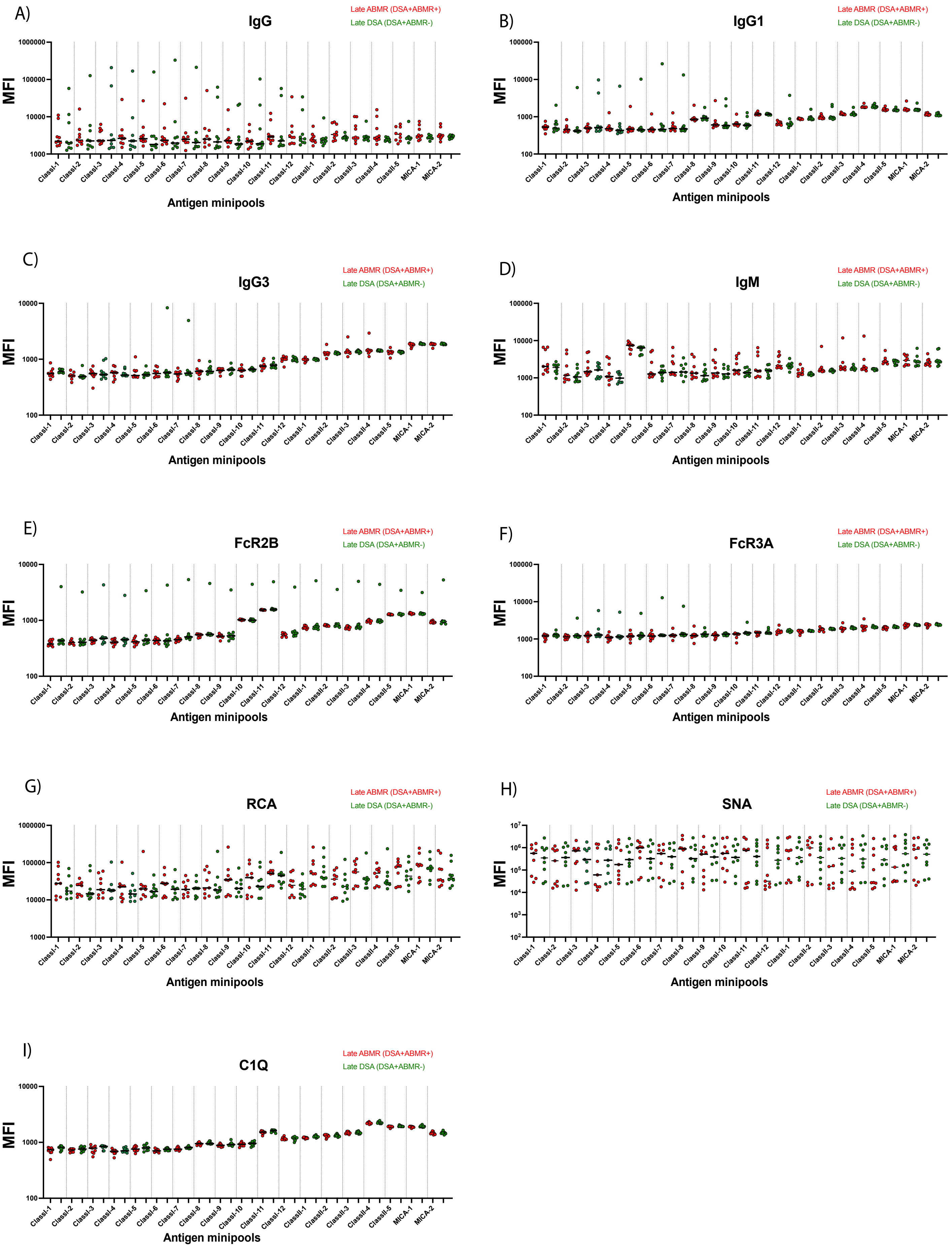
A)
IgG
Late ABMR (DSA+ABMR+)
Late DSA (DSA+ABMR-)
MFI
Antigen minipools
B)
IgG1
C)
IgG3
D)
IgM
E)
FcR2B
F)
FcR3A
G)
RCA
H)
SNA
I)
C1Q
ClassI-1
ClassI-2
ClassI-3
ClassI-4
ClassI-5
ClassI-6
ClassI-7
ClassI-8
ClassI-9
ClassI-10
ClassI-11
ClassI-12
ClassII-1
ClassII-2
ClassII-3
ClassII-4
ClassII-5
MICA-1
MICA-2
1000000
100000
10000
1000
100

fig.S6)

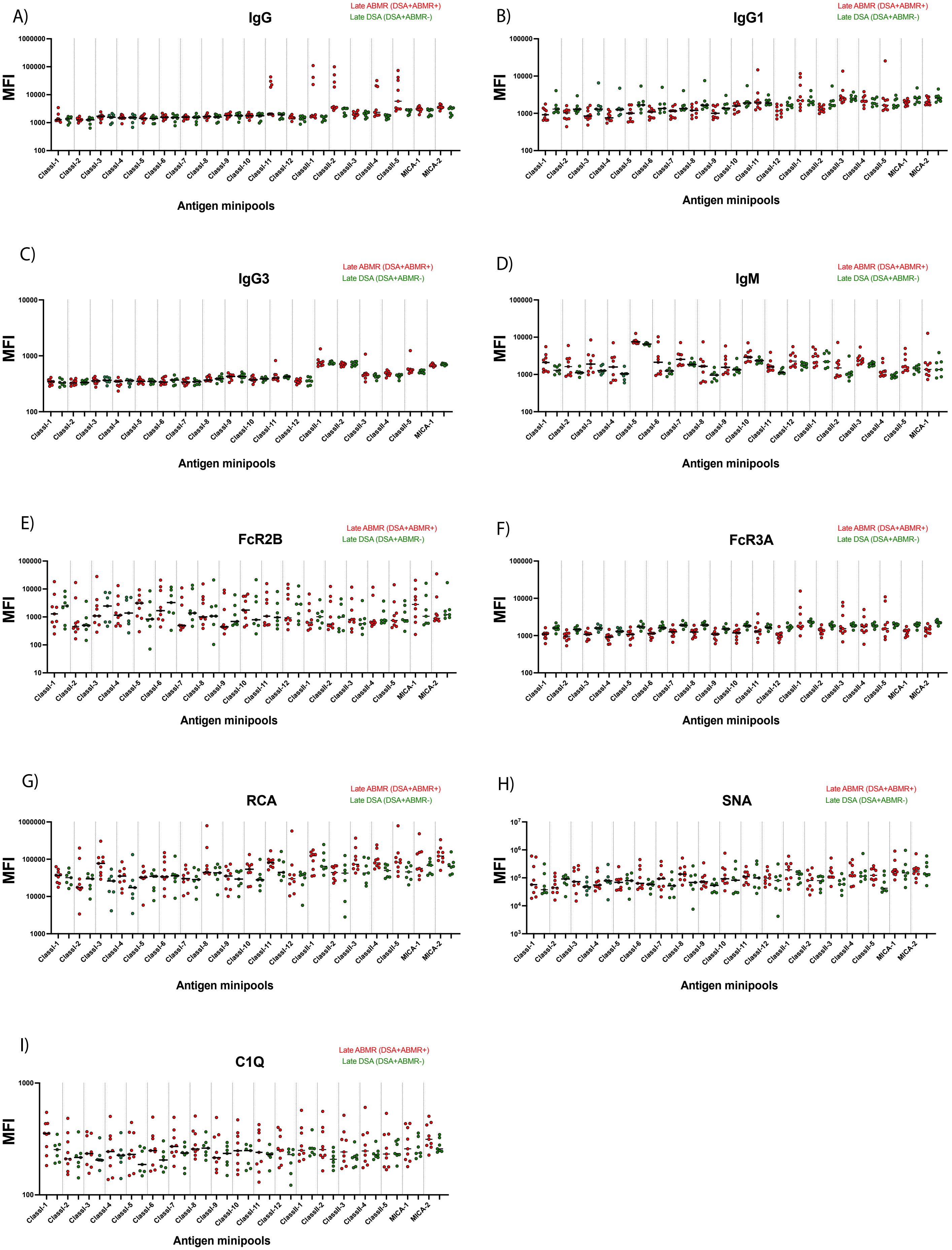

A)
IgG
Late ABMR (DSA+ABMR+)
Late DSA (DSA+ABMR-)
MFI
1000000
100000
10000
1000
100
ClassI-1
ClassI-2
ClassI-3
ClassI-4
ClassI-5
ClassI-6
ClassI-7
ClassI-8
ClassI-9
ClassI-10
ClassI-11
ClassI-12
ClassII-1
ClassII-2
ClassII-3
ClassII-4
ClassII-5
MICA-1
MICA-2
Antigen minipools
B)
IgG1
Late ABMR (DSA+ABMR+)
Late DSA (DSA+ABMR-)
MFI
100000
10000
1000
100
Antigen minipools
C)
IgG3
Late ABMR (DSA+ABMR+)
Late DSA (DSA+ABMR-)
MFI
10000
1000
100
Antigen minipools
D)
IgM
Late ABMR (DSA+ABMR+)
Late DSA (DSA+ABMR-)
MFI
100000
10000
1000
100
Antigen minipools
E)
FcR2B
Late ABMR (DSA+ABMR+)
Late DSA (DSA+ABMR-)
MFI
100000
10000
1000
100
10
Antigen minipools
F)
FcR3A
Late ABMR (DSA+ABMR+)
Late DSA (DSA+ABMR-)
MFI
100000
10000
1000
100
Antigen minipools
G)
RCA
Late ABMR (DSA+ABMR+)
Late DSA (DSA+ABMR-)
MFI
1000000
100000
10000
1000
Antigen minipools
H)
SNA
Late ABMR (DSA+ABMR+)
Late DSA (DSA+ABMR-)
MFI
$10^7$
$10^6$
$10^5$
$10^4$
$10^3$
Antigen minipools
I)
C1Q
Late ABMR (DSA+ABMR+)
Late DSA (DSA+ABMR-)
MFI
1000
100
Antigen minipools

fig.S7)

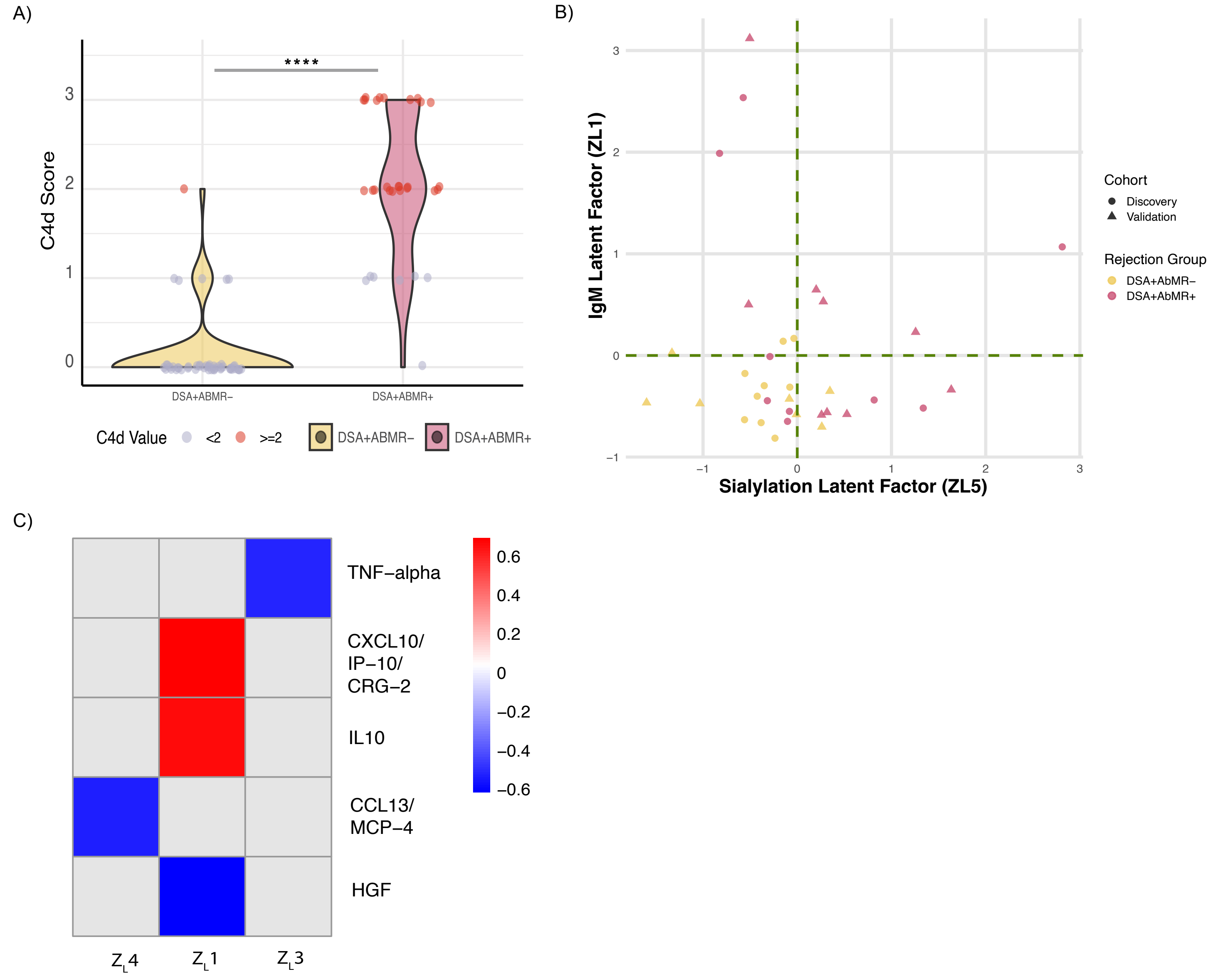

A)
****
C4d Score
0
1
2
3
DSA+ABMR−
DSA+ABMR+
C4d Value
<2
>=2
DSA+ABMR−
DSA+ABMR+
B)
IgM Latent Factor (ZL1)
Sialylation Latent Factor (ZL5)
−1
0
1
2
3
Cohort
Discovery
Validation
Rejection Group
DSA+AbMR−
DSA+AbMR+
C)
TNF−alpha
CXCL10/
IP−10/
CRG−2
IL10
CCL13/
MCP−4
HGF
0.6
0.4
0.2
0
−0.2
−0.4
−0.6
$Z_L4$
$Z_L1$
$Z_L3$

fig.S8)

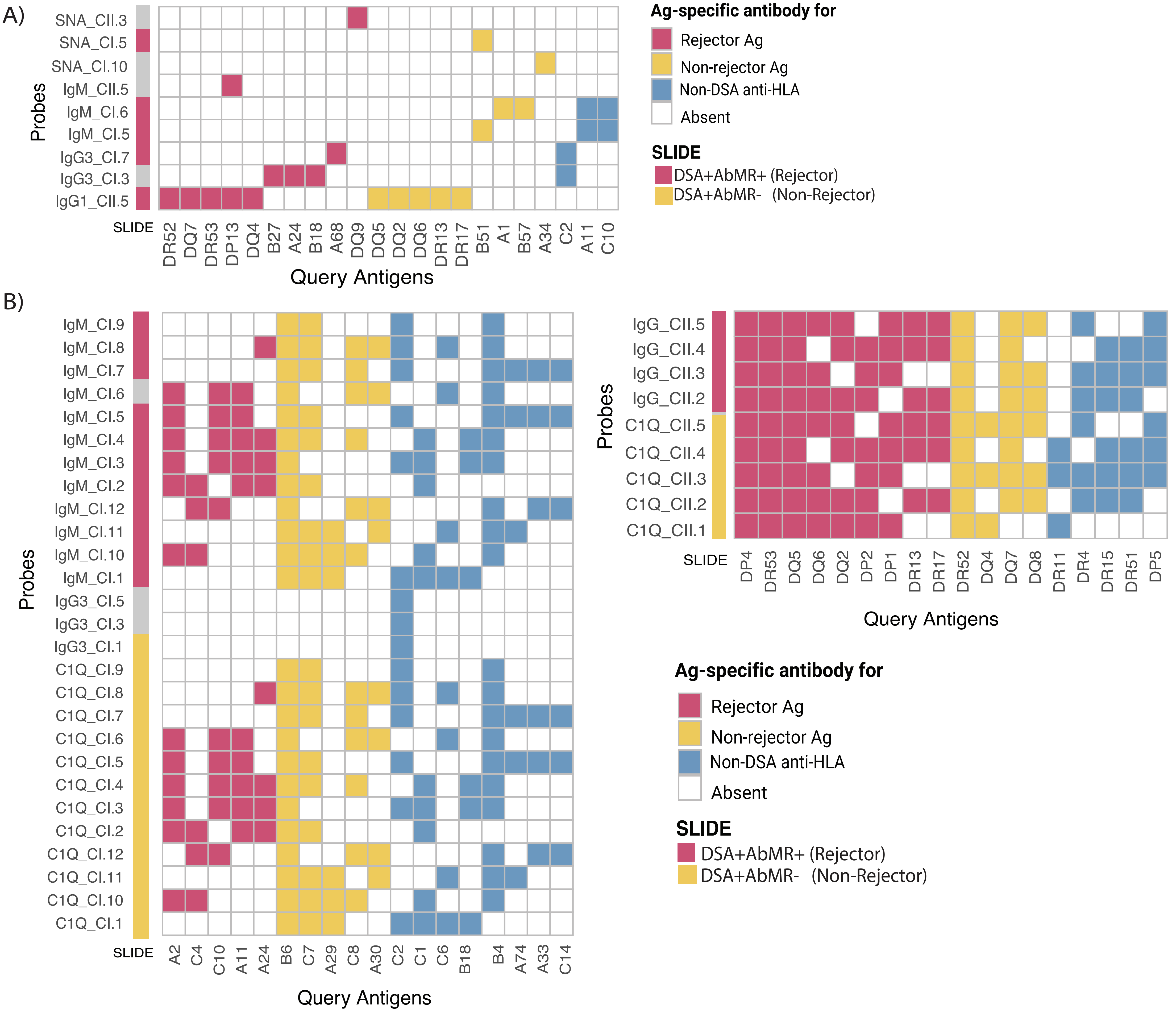
A)
Probes
SNA_CII.3
SNA_CI.5
SNA_CI.10
IgM_CII.5
IgM_CI.6
IgM_CI.5
IgG3_CI.7
IgG3_CI.3
IgG1_CII.5
SLIDE
DR52 DQ7 DR53 DP13 DQ4 B27 A24 B18 A68 DQ9 DQ5 DQ2 DQ6 DR13 DR17 B51 A1 B57 A34 C2 A11 C10
Query Antigens
Ag-specific antibody for
Rejector Ag
Non-rejector Ag
Non-DSA anti-HLA
Absent
SLIDE
DSA+AbMR+ (Rejector)
DSA+AbMR- (Non-Rejector)
B)
Probes
IgM_CI.9
IgM_CI.8
IgM_CI.7
IgM_CI.6
IgM_CI.5
IgM_CI.4
IgM_CI.3
IgM_CI.2
IgM_CI.12
IgM_CI.11
IgM_CI.10
IgM_CI.1
IgG3_CI.5
IgG3_CI.3
IgG3_CI.1
C1Q_CI.9
C1Q_CI.8
C1Q_CI.7
C1Q_CI.6
C1Q_CI.5
C1Q_CI.4
C1Q_CI.3
C1Q_CI.2
C1Q_CI.12
C1Q_CI.11
C1Q_CI.10
C1Q_CI.1
SLIDE
A2 C4 C10 A11 A24 B6 C7 A29 C8 A30 C2 C1 C6 B18 B4 A74 A33 C14
Query Antigens
Probes
IgG_CII.5
IgG_CII.4
IgG_CII.3
IgG_CII.2
C1Q_CII.5
C1Q_CII.4
C1Q_CII.3
C1Q_CII.2
C1Q_CII.1
SLIDE
DP4 DR53 DQ5 DQ6 DQ2 DP2 DP1 DR13 DR17 DR52 DQ4 DQ7 DQ8 DR11 DR4 DR15 DR51 DP5
Query Antigens
Ag-specific antibody for
Rejector Ag
Non-rejector Ag
Non-DSA anti-HLA
Absent
SLIDE
DSA+AbMR+ (Rejector)
DSA+AbMR- (Non-Rejector)

Supplementary Table 1.

**Table 1. Pittsburgh Patient Characteristics**

| Variable | N | **Overall**, N = 36[1] | **ABMR**, N = 19[1] | **DSA**, N = 17[1] | p-value[2] |
|---|---|---|---|---|---|
| **GENDER** | 36 | | | | >0.9 |
| Female | | 17 (47%) | 9 (47%) | 8 (47%) | |
| Male | | 19 (53%) | 10 (53%) | 9 (53%) | |
| **AGEATTX** | 36 | 42 (35, 59) | 40 (31, 57) | 52 (38, 61) | 0.2 |
| **Tx.number** | 36 | | | | 0.6 |
| 1 | | 24 (67%) | 12 (63%) | 12 (71%) | |
| 2 | | 9 (25%) | 6 (32%) | 3 (18%) | |
| 3 | | 3 (8.3%) | 1 (5.3%) | 2 (12%) | |
| **PRIMARY_DIAGNOSIS_AT_TRANSPLANT** | 36 | | | | 0.8 |
| ACUTE TUBULAR NECROSIS | | 1 (2.8%) | 0 (0%) | 1 (5.9%) | |
| CALCINEURIN INHIBITOR NEPHROTOXICITY | | 2 (5.6%) | 1 (5.3%) | 1 (5.9%) | |
| CHRONIC GLOMERULONEPHRITIS UNSPECIFIED | | 1 (2.8%) | 1 (5.3%) | 0 (0%) | |
| CHRONIC PYELONEPHRITIS/REFLUX NEPHROPATH | | 1 (2.8%) | 0 (0%) | 1 (5.9%) | |
| DIABETES MELLITUS - TYPE I | | 1 (2.8%) | 0 (0%) | 1 (5.9%) | |
| DIABETES MELLITUS - TYPE II | | 4 (11%) | 3 (16%) | 1 (5.9%) | |
| FOCAL GLOMERULAR SCLEROSIS (FOCAL SEGMENTAL - FSG) | | 5 (14%) | 2 (11%) | 3 (18%) | |
| HYPERTENSIVE NEPHROSCLEROSIS | | 4 (11%) | 2 (11%) | 2 (12%) | |
| IGA NEPHROPATHY | | 3 (8.3%) | 1 (5.3%) | 2 (12%) | |
| MEMBRANOUS GLOMERULONEPHRITIS | | 1 (2.8%) | 1 (5.3%) | 0 (0%) | |
| OTHER SPECIFY | | 4 (11%) | 2 (11%) | 2 (12%) | |
| POLYCYSTIC KIDNEYS | | 1 (2.8%) | 0 (0%) | 1 (5.9%) | |

[1] n (%); Median (IQR)

[2] Pearson’s Chi-squared test; Wilcoxon rank sum test; Fisher’s exact test; Wilcoxon rank sum exact test

Supplementary Table 2.

| **Variable** | **Overall**, N = 36[1] | **ABMR**, N = 19[1] | **DSA**, N = 17[1] | **p-value**[2] |
|---|---|---|---|---|
| Sample.to.Tx..mo. | 1.2 (0.9, 5.0) | 2.2 (0.9, 6.1) | 1.1 (1.0, 2.3) | 0.6 |
| Y_time | | | | 0.7 |
| Early | 18 (50%) | 9 (47%) | 9 (53%) | |
| Late | 18 (50%) | 10 (53%) | 8 (47%) | |
| FK.at.sample | | | | >0.9 |
| N | 2 (5.6%) | 1 (5.3%) | 1 (5.9%) | |
| Y | 34 (94%) | 18 (95%) | 16 (94%) | |
| MMF.at.sample | | | | 0.3 |
| N | 5 (14%) | 4 (21%) | 1 (5.9%) | |
| Y | 31 (86%) | 15 (79%) | 16 (94%) | |
| PREDNISONE.at.sample | | | | 0.040 |
| N | 4 (11%) | 0 (0%) | 4 (24%) | |
| Y | 32 (89%) | 19 (100%) | 13 (76%) | |

[1] Median (IQR); n (%)

[2] Wilcoxon rank sum test; Pearson's Chi-squared test; Fisher's exact test

| Toronto Transplant Cohort (n=32) | | |
|---|---|---|
| **Demographics** | | |
| Sex, n male, % | 24 | 75% |
| Age at tx, mean yrs, std | 45 | 17.9 |
| Age at bx, mean yrs, std | 50 | 15.5 |
| *Recipient self-reported race (n, %)* | | |
| Asian | 5 | 16% |
| White/Middle Eastern | 18 | 56% |
| Not Specified | 9 | 28% |
| *Recipient cause of ESKD (n, %)* | | |
| Diabetes mellitus | 6 | 19% |
| Focal and segmental glomerulosclerosis | 5 | 16% |
| IgA Nephropathy | 3 | 9% |
| Polycystic kidney disease | 2 | 6% |
| Vasculitis | 2 | 6% |
| Other | 11 | 34% |
| Unknown | 3 | 9% |
| *Recipient co-morbidities (n, %)* | | |
| Hypertension | 30 | 94% |
| Diabetes | 12 | 38% |
| Autoimmune disease | 7 | 22% |
| Coronary heart disease | 8 | 25% |
| Non-skin cancer | 7 | 22% |
| *Donor-specific antibodies (DSA)* | | |
| n DSA, mean, std, | 1.1 | 0.4 |
| Anti-HLA Class I DSA, n patients, % | 11 | 34% |
| Anti-HLA Class I DSA, mean Abs, std | 0.3 | 0.5 |
| Anti-HLA Class II DSA, n patients, % | 24 | 75% |
| Anti-HLA Class II DSA, mean Abs, std | 0.8 | 0.5 |
| **Transplant parameters/outcomes** | | |
| Transplanted organ, n kidney only, % | 28 | 88% |
| Renal transplant number, n first tx, % | 29 | 91% |
| Induction agent, n anti-thymoglobulin, % | 22 | 69% |
| Treatment for HLA desensitization, n. % | 2 | 6% |
| Time Tx-Bx, n days, std | 1942 | 2625 |
| Donor type, n living, % | 14 | 44% |
| Serum creatinine at bx, mean µmol/L, std | 268 | 389 |
| Delayed graft function, n, % | 6 | 19% |
| Death-censored graft loss, n, % | 9 | 28% |
| Death, n, % | 4 | 13% |
| *Maintenance immunosuppression at Bx (n, %)* | | |
| Tacrolimus | 27 | 84% |
| Cyclosporine | 4 | 13% |
| Cellcept/MMF | 26 | 81% |
| Azathioprine | 2 | 6% |
| Other | 2 | 6% |
| Prednisone | 31 | 97% |
| *Treatment for rejection at Bx (n, %)* | | |
| Steroid | 10 | 31% |
| Increased immunosuppression | 5 | 16% |
| IVIG | 10 | 31% |
| PLEX | 7 | 22% |
| None | 18 | 56% |
| *Banff rejection category (n, %)* | | |
| NR | 15 | 47% |
| Active AMR | 2 | 6% |
| Chronic AMR | 12 | 38% |
| Mixed AMR | 2 | 6% |
| TCMR | 1 | 3% |
| *Histology (mean, max)* | | |
| % sclerosed glomeruli | 14 | 66 |
| glomerulitis (g) | 1 | 3 |
| mesangial matrix expansion (mm) | 0 | 2 |
| transplant glomerulopathy (cg) | 1 | 3 |
| interstitial inflammation (i) | 0 | 1 |
| tubulitis (t) | 0 | 2 |
| peritubular capillaritis (ptc) | 1 | 3 |
| interstitial fibrosis (ci) | 1 | 3 |
| tubular atrophy (ct) | 1 | 3 |
| intimal arteritis (v) | 0 | 0 |
| arteriolar hyalinosis (ah) | 1 | 3 |
| vascular fibrous intimal thickening (cv) | 1 | 3 |
| C4d deposition (c4d) | 0 | 2 |
